# Driving Under the Influence of Alcohol in People with Major Depressive Episodes and Alcohol Use Disorder

**DOI:** 10.1101/2023.09.20.23295856

**Authors:** Ji-Yeun Park, Brent M. Peterson, Jinsil Kim, Thushara Galbadage

**Author notes:** Corresponding Authors Ji-Yeun Park, PhD, Thushara Galbadage, PhD, MPH, Department of Kinesiology and Public Health, 13800 Biola Ave, La Mirada, CA 90639.

## Abstract

**Objectives:** Alcohol use disorder (AUD) and depression are the most commonly reported psychiatric comorbid conditions. We examined trends in the past-year prevalence of driving under the influence of alcohol (DUIA) among people with major depressive episodes (MDE), AUD, or both in the United States.

**Methods:** We analyzed 543,573 individuals aged 18 years or older from the 2005-2019 National Surveys on Drug Use and Health (NSDUH). Multivariate logistic regression models were applied to examine the adjusted past-year prevalence of DUIA. To assess trends in DUIA over time, annual average percent change (AAPC) was calculated.

**Results:** From 2005 to 2019, DUIA prevalence among US adults with MDE declined significantly from 18.1% to 9.4% (AAPC = -4.9). Decreasing trends in DUIA were also observed among those with AUD (from 55.4% to 37.8%, AAPC = -3.0) and among those with co-occurring MDE and AUD (from 58.3% to 38.8%, AAPC = -3.1). Compared to those with no AUD or MDE (from 8.42% to 3.99%, AAPC = -5.6), individuals with AUD and those with co-occurring MDE and AUD had significantly lower AAPCs. There was no significant AAPC difference between those with MDE and those with no AUD or MDE. Regarding sociodemographic characteristics, younger adults aged 18-34 exhibited the lowest AAPC among all diagnostic groups.

**Conclusions:** From 2005 to 2019, DUIA prevalence declined significantly with varying rates of decrease across different diagnostic groups. Focused public health efforts are needed to engage high-risk groups that have shown a tendency toward less expedient reductions in DUIA.

## 1. INTRODUCTION

The prevalence of driving under the influence of alcohol (DUIA) among the overall US population decreased from 12.1% in 2016 to 10.7% in 2020 ^1^. Despite these decreasing trends, however, DUIA remains an ongoing public health concern in the US. The US Department of Transportation has reported that approximately 11,654 people died from drunk-driving-related crashes in 2020, representing a 14% increase from 2019 ^2^. This equates to one person dying from alcohol-related accidents every 45 minutes. Furthermore, economic losses from alcohol-related crashes accounted for $68.9 billion in 2019 ^3^.

DUIA is influenced by many risk factors. The risk of DUIA varies across sociodemographic subgroups. Males, white individuals, young adults, and those with high socioeconomic status are at elevated risk of DUIA ^4-6^. Alcohol use disorder (AUD) is the primary risk factor associated with DUIA ^7,8^. Previous studies have shown that a significant proportion of both first and repeated DUIA offenders had AUD ^9,10^. Mental health problems are another major contributing factor to DUIA ^9-12^. Several studies have demonstrated a higher prevalence of mental health disorders, such as depression, among those who reported DUIA compared to the general population ^13-16^.

While there have been numerous studies on predictors associated with DUIA, particularly behavioral risk factors like depression and ADU, few studies have examined the trends in DUIA among individuals with various comorbidities, including MDE, AUD, or both. Mental disorders often manifest alongside AUD, with depressive disorders being the most common comorbidity ^17^. Given the high rates of comorbidity between these disorders, it is crucial to understand the overall proportion of people who reported DUIA across different diagnostic groups ^18^. Furthermore, a better understanding of how DUIA prevalence has changed over time across different diagnostic groups will provide valuable insight into the development of DUIA intervention programs targeting those at higher risk of DUIA. It is also important to consider the role of sociodemographic factors when looking at the trends in DUIA among people with MDE, AUD, or both. The prevalence of self-reported DUIA has been shown to differ across various sociodemographic subgroups, including age, sex, and annual household income ^5,7^. Similarly, differences are documented in sociodemographic subgroups in individuals experiencing MDE and/or AUD ^19-22^. Accordingly, the current study was designed to address these significant research gaps by analyzing nationally representative data from 2005 - 2019 National Survey on Drug Use and Health (NSDUH). The goals of this study were to (1) assess changes in trends in DUIA prevalence across different diagnostic groups (MDE, AUD, or both) and (2) further explore the sociodemographic differences among individuals with AUD, MDE, or both who reported DUIA. This study provides new information on the average annual percent change (AAPC) in the self-reported DUIA prevalence by sociodemographic and diagnostic conditions, which has not been addressed in previous research. This information is useful for developing tailored interventions to reduce DUIA prevalence.

## 2. METHODS

### 2.1. Data

The NSDUH provides nationally representative data on the use of tobacco, alcohol, legal and illegal substances, and mental health problems among US civilian, noninstitutionalized populations. Respondents were interviewed using a combination of computer-assisted interviewing and audio computer-assisted self-interviewing (ACASI). The use of ACASI provides respondents with a highly private and confidential way to respond more honestly to questions about illicit drug use and other sensitive behaviors. We analyzed data from individuals aged 18 years or older who participated in the 2005 - 2019 NSDUH. While the NSDUH data are obtained from human subjects, the secondary dataset utilized in this study is publicly available and de-identified, which has been verified as not meeting the criteria necessary for review and approval as human subject research.

### 2.2. Measures

The survey assessed DUIA behaviors by asking the question ‘During the past 12 months, have you driven a vehicle while you were under the influence of alcohol?’ Responses were categorized as a binary variable (yes or no). The NSUDH assessed the past-year MDE and AUD using diagnostic criteria specified in the fourth edition of the Diagnostic and Statistical Manual of Mental Disorders (DSM-IV) ^23^. Respondents were considered to have past-year MDE if they experienced at least five out of the nine symptoms of MDE within 2 weeks or longer, based on the DSM-IV criteria. Those who met the DSM-IV criteria for AUD were classified as having past-year AUD. Additionally, self-reported sociodemographic characteristics, such as sex, age, education, annual household income, marital status, and metropolitan status, were assessed. We grouped race/ethnicity into four subgroups (NH-White, NH-Black, Hispanic, and Others).

### 2.3. Statistical Analysis

In 2015, the NSDUH made partial changes to the questionnaire that affected respondents eligible to be asked questions about DUIA. Additional changes were made in 2016 by introducing questions about driving under the influence of specific drugs, such as marijuana, pain relievers, and cocaine. Due to these changes in the survey design, DUIA established new baseline trends in 2016. Therefore, we used NSDUH 2016 - 2019 to calculate the prevalence ratio (PR) with confidence intervals (CIs), stratified by sex, age, race/ethnicity, education, income, marital status, metropolitan status, past-year MDE, and past-year AUD.

To assess the long-term trends of DUIA over time, we calculated the adjusted past-year prevalence of DUIA from 2005 to 2019. The prevalence estimate for each year was adjusted for all covariates, including sex, age, education, annual household income, marital status, and metropolitan status. Long-term trends of DUIA were examined by past-year MDE, AUD, or both. Additionally, we conducted subgroup analyses by selected sociodemographic characteristics and then calculated AAPC to compare the magnitude of change in DUIA rates across various subpopulations. Joinpoint Trend Analysis software version 4.9.1.0 (National Cancer Institute, 2023) was utilized to estimate AAPC values. For all other analyses, STATA version 17 (College Station, TX) was employed to determine the NSDUH’s sampling weights. All data used in this study and analysis results have been placed in a public repository. This dataset can be accessed at https://doi.org/10.7910/DVN/W2VPKC.

## 3. RESULTS

### 3.1. Prevalence of DUIA by sociodemographic and diagnostic conditions

Table 1 shows the past-year prevalence for 2016 - 2019 and the unadjusted/adjusted PRs of DUIA. In the overall population, the unadjusted prevalence of DUIA was 8.3%. The past-year prevalence of self-reported DUIA was significantly higher in men (10.8% [95% CI, 10.6% - 11.1%]), younger adults aged 26 – 34 (11.7% [95% CI, 11.2% - 12.2%]), NH-White individuals (10.0% [95% CI, 9.7% - 10.3%]), those with higher levels of education (some college but no degree; 8.9% [95% CI, 8.6% - 9.2%], college degree; 8.3% [95% CI, 8.1% - 8.5%]), those with family incomes over $75,000 (11.2% [95% CI, 10.9% - 11.5%]), individuals who never married (10.7% [95% CI, 10.4% - 11.1%]), and those residing in large and small metropolitan areas (8.5% [95% CI, 8.2% - 8.8%] and 8.5% [95% CI, 8.2% - 8.9%], respectively). Compared to those without MDE (8.0% [95% CI, 7.8% - 8.2%]), individuals with MDE had a significantly higher prevalence of DUIA (13.1% [95% CI, 12.4% - 13.9%]). Past-year prevalence of DUIA was markedly higher in people with AUD (46.9% [95% CI, 45.4% - 48.3%]) than in those without AUD (6.0 [95% CI, 5.8% - 6.2%]).

**Table 1.**
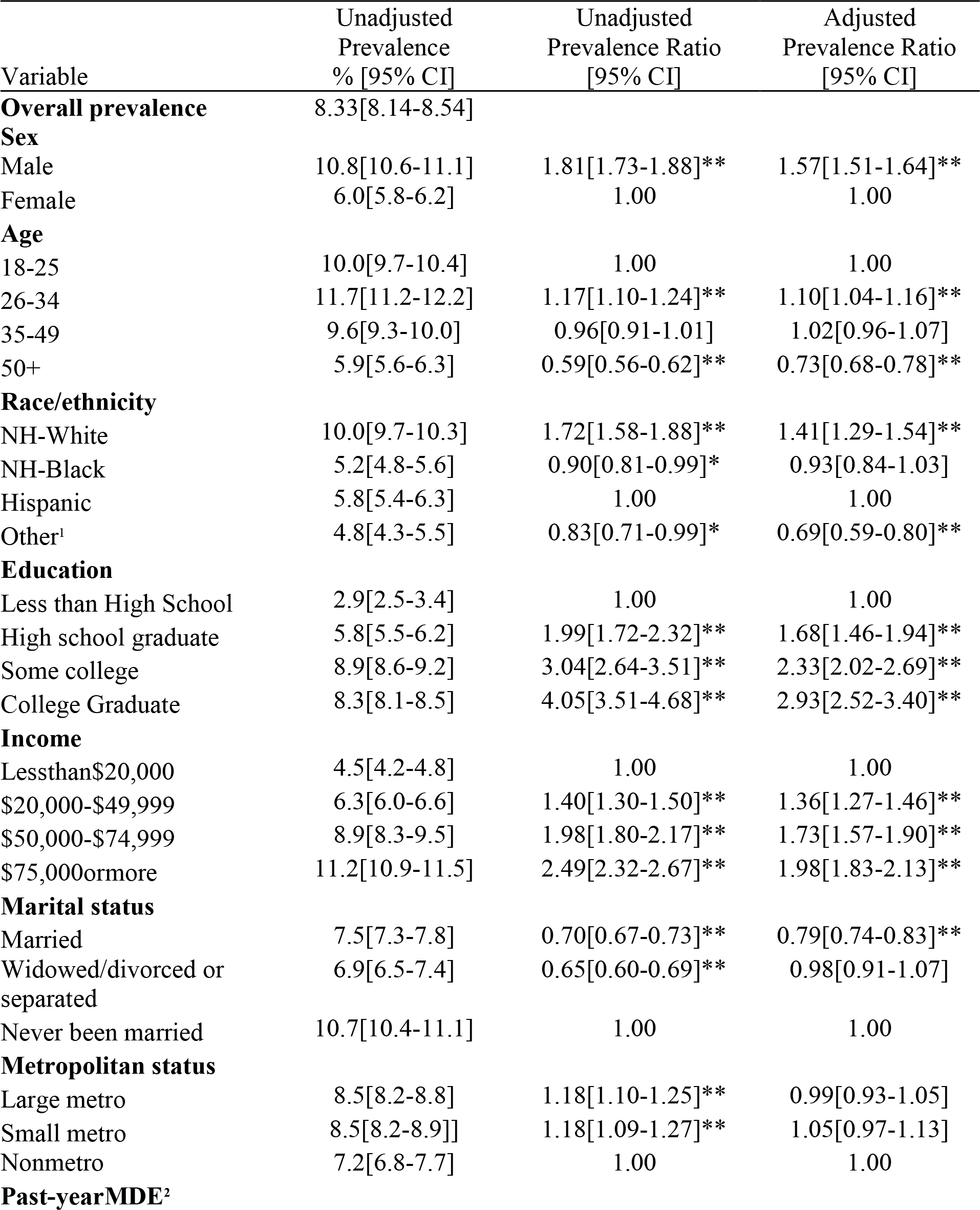
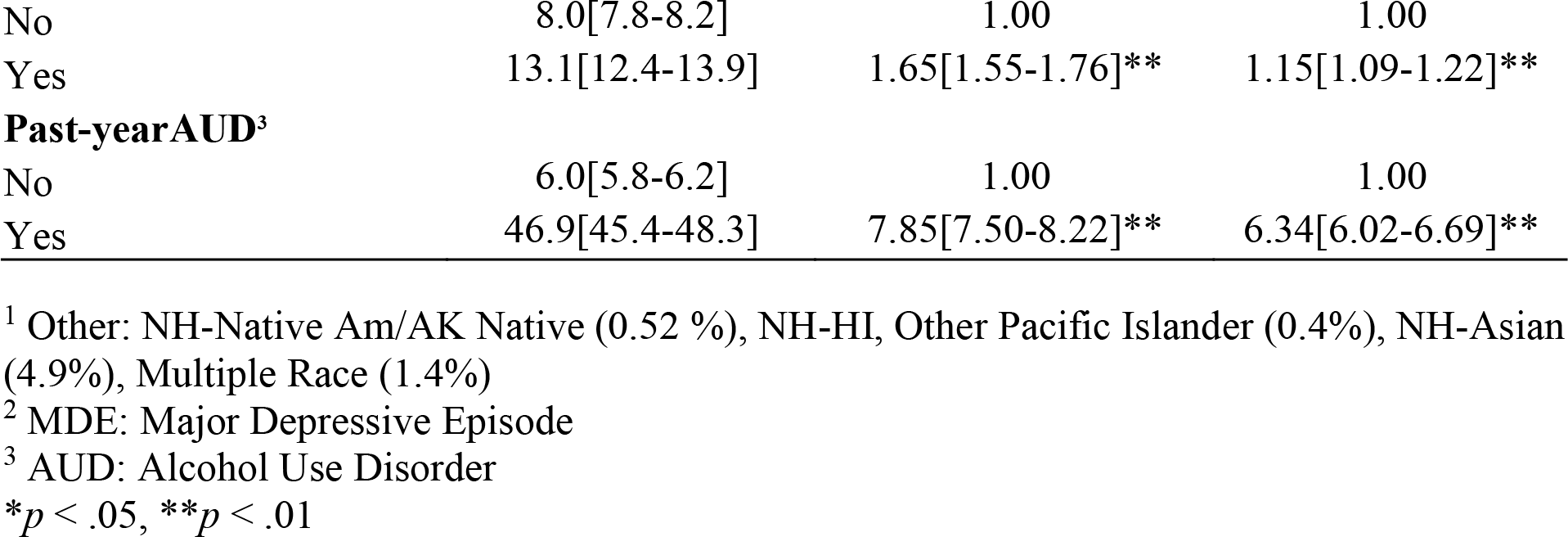
Prevalence of driving under the influence of alcohol by sociodemographic and psychiatric characteristics, NSDUH 2016 - 2019.

Married individuals had decreased unadjusted and adjusted odds ratio of DUIA prevalence compared to never-married individuals. Men, young adults aged 26 – 34, NH-White individuals, those with higher levels of education, and those with higher household income had increased unadjusted and adjusted odds ratios of DUIA prevalence compared to their respective counterparts. People with MDE had significantly increased odds of DUIA (unadjusted PR = 1.7, 95% CI, 1.6 - 1.8; adjusted PR = 1.2, 95% CI, 1.1 - 1.2, all *p* < .01) than people without MDE. Individuals with AUD had the highest strength of association with DUIA (unadjusted PR = 7.9, 95% CI, 7.5 - 8.2; adjusted PR = 6.3, 95% CI, 6.0 - 6.7, all *p* < .01).

### 3.2. Trends in the Prevalence of Past-year DUIA

#### 3.2.1. By diagnostic conditions (MDE, AUD, or both)

From 2005 to 2019, there was a significant reduction in self-reported DUIA prevalence among people with MDE, AUD, or both. The adjusted past-year prevalence of DUIA declined from 18.1% to 9.4% among adults with past-year MDE for an AAPC of -4.9 (95% CI, -6.3 to -3.6; *p* < .001; Figure 1 and Supplemental Table 1). The adjusted past-year prevalence of DUIA declined from 55.4% to 37.8% among adults with past-year AUD for an AAPC of -3.0 (95% CI, -3.8 to - 2.1; *p* < .001; Figure 1 and Supplemental Table 1). The adjusted past-year prevalence of DUIA declined from 58.3% to 38.8% among adults with co-occurring MDE and AUD for an AAPC of -3.1 (95% CI, -3.8 to -2.4; *p* < .001; Figure 1 and Supplemental Table 1). Despite the declining trend in DUIA prevalence among US adults with MDE, AUD, or both, the prevalence was significantly higher during 2005 – 2019 than among those without MDE (11.3% to 5.7% for an AAPC of -5.2 [95% CI, -6.7 to -3.8]), AUD (8.5% to 4.4% for an AAPC of -5.2 [95% CI, -6.7 to -3.7]), or both (8.4% to 4.0% for an AAPC of -5.6 [95% CI, -7.0 to -4.2]) (all *p* < .001; Figure 1 and Supplemental Table 1).

**Figure 1.**
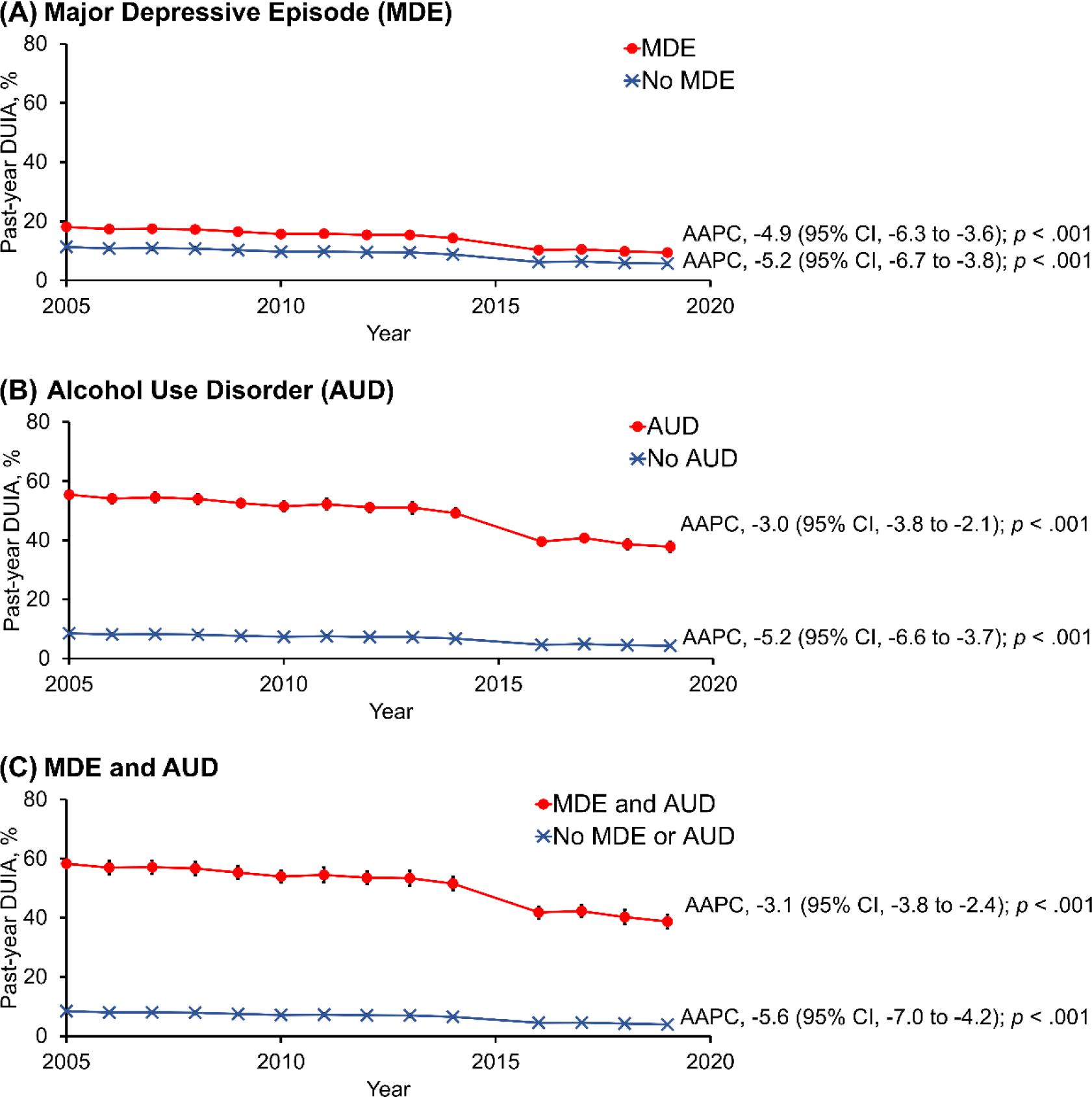
Trends in adjusted past-year prevalence of driving under the influence of alcohol from 2005 to 2019, by MDE and AUD status. (A) Major Depressive Episode (MDE), (B) Alcohol Use Disorder (AUD), and (C) MDE and AUD. Average annual percent changes (AAPC) and their 95% confidence intervals (95% CI) are indicated for each of the trend graphs. Error bars represent the standard errors. Each *p*-value indicates changes during 2005-2019. Due to the changes in the questionnaire that asked questions about DUIA, the 2015 data on DUIA were not available from the NSDUH 2002-2019 public use data file.

#### 3.2.2. By diagnostic and sociodemographic conditions

During 2005 - 2019, adjusted past-year DUIA prevalence declined significantly for all examined sociodemographic subgroups (age, sex, race/ethnicity, education, income, marital status, and metropolitan status) with MDE (all *p* < .001; Figure 2, Supplemental Figure 1, and Supplemental Tables 2-8). Among those with MDE, the highest AAPC was found in adults aged 50 or older (-5.3 [95% CI, -6.7 to -3.9]; *p* < .001; Supplemental Table 3D), other racial/ethnic groups (-5.3 [95% CI, -6.6 to -4.0]; *p* < .001; Supplemental Table 4D), and those without high school diploma (-5.3 [95% CI, -6.7 to -3.9]; *p* < .001; Supplemental Table 6A). The lowest AAPC was found in adults aged 18 - 25 years (-4.6 [95% CI, -5.7 to -3.4]; *p* < .001; Supplemental Table 3A) and adults aged 26 - 34 years (-4.6 [95% CI, -5.8 to -3.4]; *p* < .001; Supplemental Table 3B). Adjusted past-year DUIA prevalence also declined significantly for all examined sociodemographic subgroups with AUD (all *p* < .001; Figure 2, Supplemental Figure 1, and Supplemental Tables 2-8). Among those with AUD, the highest AAPC was found in other racial/ethnic groups (-3.8 [95% CI, -4.8 to -2.7]; *p* < .001; Supplemental Table 4D) and those without high school diploma (-3.8 [95% CI, -4.9 to -2.8]; *p* < .001; Supplemental Table 6A). The lowest AAPC was found in adults aged 26 - 34 years (-2.3 [95% CI, -3.0 to -1.7]; *p* < .001; Supplemental Table 3B).

**Figure 2.**
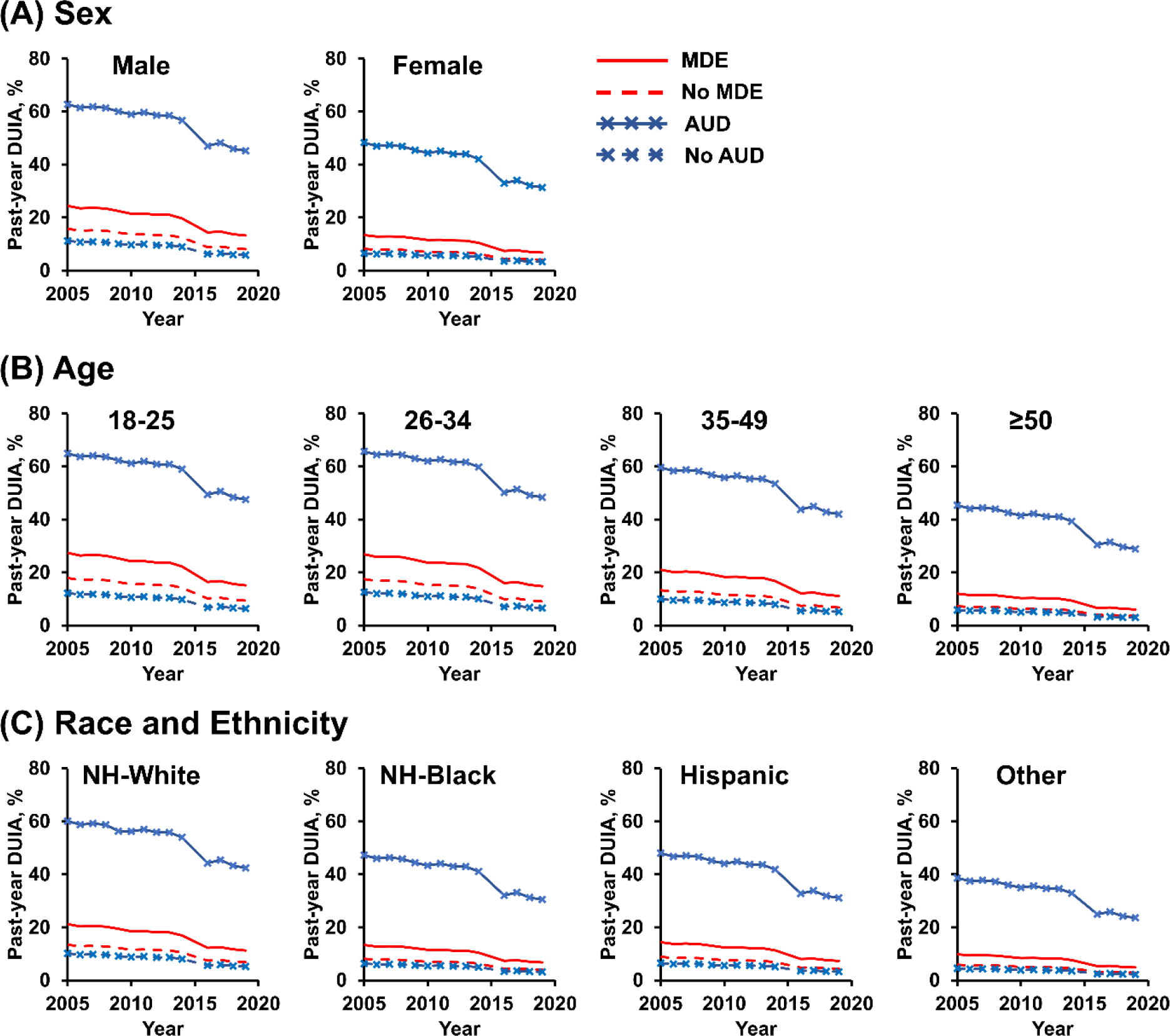
Trends in adjusted past-year prevalence of self-reported driving under the influence of alcohol among US adults with or without MDE or AUD, by demographic characteristics. (A) Past-year prevalence of DUIA stratified according to sex. (B) Past-year prevalence of DUIA stratified according to age. (C) Past-year prevalence of DUIA stratified according to race and ethnicity. The figure legend on the top right corner applies to all sub-graphs (A to C). Due to the changes in the questionnaire that asked questions about DUIA, the 2015 data on DUIA were not available from the NSDUH 2002-2019 public use data file.

Additionally, adjusted past-year DUIA prevalence declined significantly for all examined sociodemographic subgroups with co-occurring MDE and AUD (all *p* < .001; Supplemental Figures 23 and Supplemental Tables 2-8). Among those with co-occurring MDE and AUD, the highest AAPC was found in other racial/ethnic groups and those without high school diplomas (-4.0 [95% CI, -4.9 to -3.0]; *p* < .001, respectively; Supplemental Tables 4D and 6A). The lowest AAPC was found in adults aged 18 - 25 years and adults aged 26 - 34 years (-2.4 [95% CI, -3.0 to -1.9]; *p* < .001, respectively; Supplemental Table 3A and 3B).

## 4. DISCUSSION

In this study, we aimed to assess trends in DUIA prevalence among individuals with MDE, AUD, or both, using a nationally representative sample. Our findings revealed that between 2005 and 2019, the overall prevalence of DUIA among people with MDE, AUD, or both declined significantly. Nevertheless, despite these decreasing trends, DUIA prevalence rates in these populations remain considerably high.

As expected, people with co-occurring MDE and AUD had a significantly higher prevalence of DUIA than those with MDE only. Furthermore, those with AUD only also had a significantly higher prevalence of DUIA than individuals with MDE only. However, DUIA prevalence rates were largely similar among individuals with comorbid MDE and AUD and those with AUD only. These findings support previous evidence suggesting that MDE alone was modestly associated with DUIA, while AUD itself was strongly associated with DUIA ^15^. Although results from this study were not able to elucidate the extent to which depression contributes to the association between AUD and DUIA, findings reported herein indicate that depression may have little or no influence on DUIA among individuals with AUD.

One of the most striking findings of this study is that the overall rate of decline in DUIA was significantly different across diagnostic groups. Specifically, compared to those with no AUD or MDE, individuals with AUD and those with co-occurring MDE and AUD had a significantly lower rate of decline. These findings emphasize the urgent need for interventions to accelerate progress in DUIA prevention within these high-risk population segments. Notably, there was no significant difference in the rate of decline between those with MDE and those with no AUD or MDE. Such findings suggest that while depression is theoretically an important predictor of DUIA, it may indirectly contribute to DUIA through problematic patterns of alcohol use. A previous study using path analysis showed that depression was associated with DUIA frequency, although such an association is mediated by AUD ^15^.

The rate of decline in DUIA among people with MDE, AUD, or both varied significantly across sociodemographic characteristics. Younger adults consistently had the lowest AAPC regardless of their diagnostic status. Specifically, those aged 26 – 34 with AUD and those aged 18 - 34 with comorbid MDE and AUD had a significantly lower rate of decline compared to other subgroups. These findings suggest that DUIA patterns in people with AUD and MDE may be influenced by age. For example, one possible explanation is that young adults tend to have a lower risk perception of DUIA than older adults, which consequently, may increase the risk of DUIA ^24^.

These findings have implications for reducing DUIA prevalence. First, DUIA prevention efforts should prioritize individuals with AUD and those with co-occurring MDE and AUD because they are not only at significantly greater risk for DUIA but also exhibit a slower rate of decline. In clinical practice, treatment specifically tailored to address both MDE and AUD may play a crucial role in reducing DUIA. Second, age differences in DUIA among individuals with AUD and MDE should be considered when designing intervention programs. Tailoring DUIA prevention to younger adults diagnosed with AUD and MDE may significantly contribute to further reducing DUIA prevalence.

This study has several limitations. The NSDUH survey data are cross-sectional, which means that we cannot infer any causal relationships. Second, the NSDUH data collected are self-reported, making them susceptible to recall, social desirability, and self-reporting biases. Additionally, MDE is the only non-substance use mental health disorder measure in the NSDUH survey. Therefore, any potential associations DUIA has with other psychotic disorders and other mental disorders cannot be determined by using this dataset. Furthermore, the NSDUH survey excludes people experiencing homelessness not living in shelters, and institutionalized (e.g., jail or prison) populations. These populations often experience higher rates of AUD and mental illness which may result in an underestimation of the prevalence of MDE and AUD in this study ^25^.

## 5. CONCLUSIONS

Our study findings contribute to the growing body of evidence that, while DUIA prevalence significantly declined among people with AUD, MDE, or both, the magnitude of change varied across diagnostic groups. Furthermore, younger adults with AUD and those with co-occurring AUD and MDE exhibited significantly lower AAPC. Therefore, when developing effective interventions to further reduce DUIA, it is crucial to take into consideration an individual’s comorbidity status and age. Prevention efforts could be targeted at simultaneously treating depression and AUD symptoms and providing education to change attitudes and risk perceptions toward DUIA, especially among younger adults.

## Data Availability

All data produced in the present study are available upon reasonable request to the authors

https://doi.org/10.7910/DVN/W2VPKC

