## Supplementary Information for "Driving Under the Influence of Alcohol in People with Major Depressive Episodes and Alcohol Use Disorder"

**SUPPLEMENTAL INFORMATION**

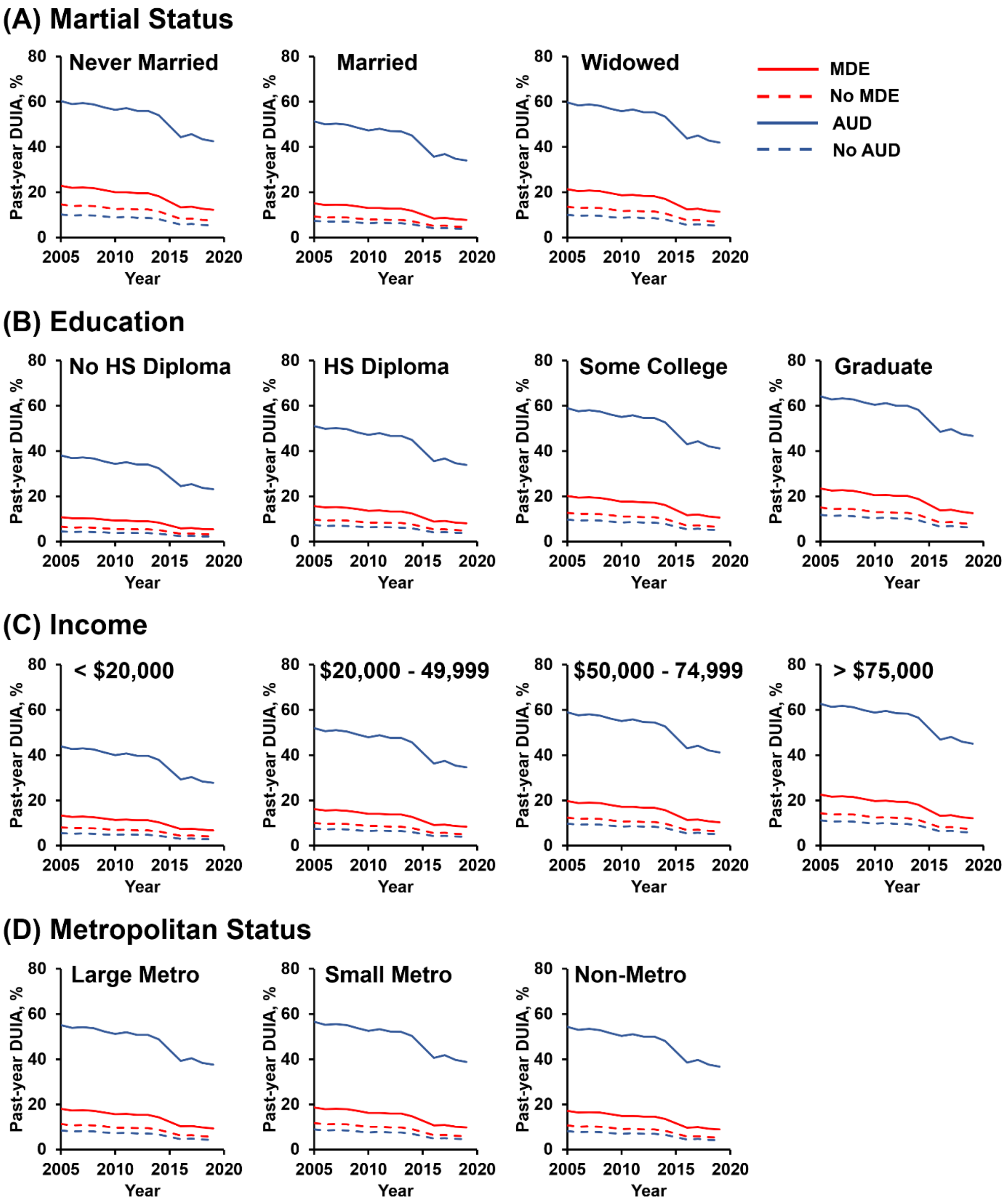

**Supplemental Figure 1. Trends in adjusted past-year prevalence of self-reported driving under the influence of alcohol among US adults with or without MDE or AUD, by demographic and socioeconomic characteristics.** (A) Past-year prevalence of DUIA stratified according to marital status. (B) Past-year prevalence of DUIA stratified according to education. (C) Past-year prevalence of DUIA stratified according to income. (D) Past-year prevalence of DUIA stratified according to metropolitan status. The figure legend on the top right corner applies to all sub-graphs (A to D). Due to the changes in the questionnaire that asked questions about DUIA, the 2015 data on DUIA were not available from the NSDUH 2002-2019 public use data file.

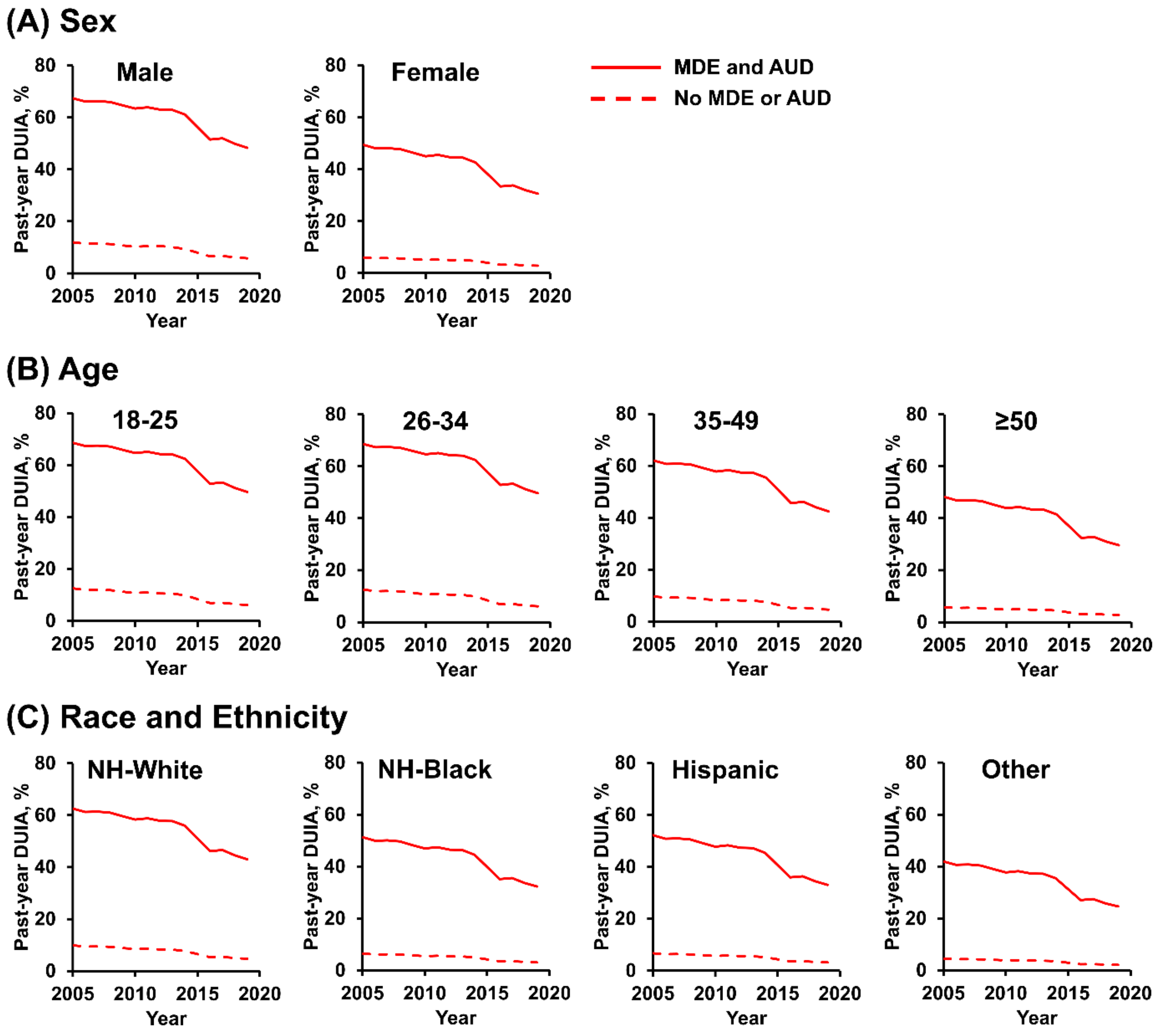


**Supplemental Figure 2. Trends in adjusted past-year prevalence of self-reported driving under the influence of alcohol among US adults with both MDE and AUD or neither, by demographic characteristics.** (A) Past-year prevalence of DUIA stratified according to sex. (B) Past-year prevalence of DUIA stratified according to age. (C) Past-year prevalence of DUIA stratified according to race and ethnicity. The figure legend on the top right corner applies to all sub-graphs (A to C). Due to the changes in the questionnaire that asked questions about DUIA, the 2015 data on DUIA were not available from the NSDUH 2002-2019 public use data file.

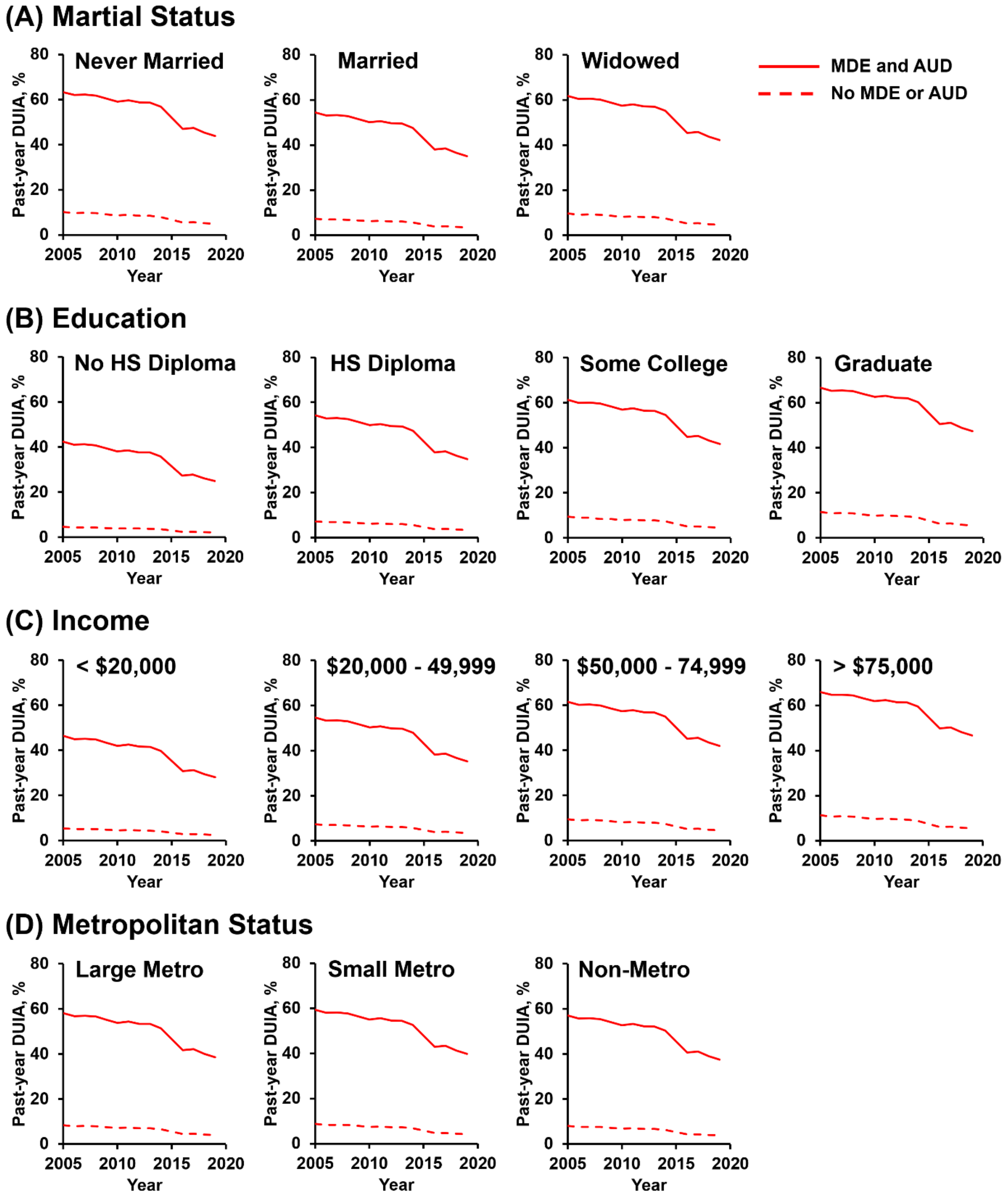


**Supplemental Figure 3**. **Trends in adjusted past-year prevalence of self-reported driving under the influence of alcohol among US adults with both MDE and AUD or neither, by demographic and socioeconomic characteristics.** (A) Past-year prevalence of DUIA stratified according to marital status. (B) Past-year prevalence of DUIA stratified according to education. (C) Past-year prevalence of DUIA stratified according to income. (D) Past-year prevalence of DUIA stratified according to metropolitan status. The figure legend on the top right corner applies to all sub-graphs (A to D). Due to the changes in the questionnaire that asked questions about DUIA, the 2015 data on DUIA were not available from the NSDUH 2002-2019 public use data file.

**Supplemental Table 1.** Trends in past-year adjusted prevalence of driving under the influence of alcohol (DUIA) and 95% CI by past-year major depressive episode (MDE) and alcohol use disorder (AUD), from 2005 to 2019.

| Year | MDE[1] | No MDE[2] | AUD[3] | No AUD[4] | MDE and AUD[5] | No MDE or AUD[6] |
|---|---|---|---|---|---|---|
| 2005 | 18.1 [17.24-18.96] | 11.32 [10.82-11.82] | 55.41 [53.91-56.91] | 8.53 [8.1-8.95] | 58.33 [56.34-60.32] | 8.42 [8.02-8.82] |
| 2006 | 17.33 [16.47-18.2] | 10.8 [10.34-11.27] | 54.09 [52.56-55.62] | 8.12 [7.73-8.52] | 57 [54.79-59.21] | 8.01 [7.63-8.38] |
| 2007 | 17.5 [16.61-18.39] | 10.91 [10.43-11.4] | 54.49 [52.8-56.17] | 8.24 [7.79-8.69] | 57.13 [54.84-59.42] | 8.05 [7.64-8.46] |
| 2008 | 17.27 [16.36-18.17] | 10.76 [10.23-11.28] | 53.98 [52.26-55.69] | 8.09 [7.66-8.52] | 56.71 [54.41-59.02] | 7.92 [7.51-8.33] |
| 2009 | 16.52 [15.72-17.32] | 10.26 [9.8-10.71] | 52.56 [50.94-54.19] | 7.68 [7.29-8.06] | 55.33 [53.23-57.42] | 7.52 [7.16-7.89] |
| 2010 | 15.71 [14.86-16.57] | 9.72 [9.27-10.17] | 51.44 [49.81-53.07] | 7.36 [6.93-7.79] | 53.99 [51.9-56.08] | 7.15 [6.78-7.53] |
| 2011 | 15.79 [14.95-16.62] | 9.77 [9.3-10.24] | 52.21 [50.46-53.96] | 7.58 [7.15-8] | 54.51 [51.96-57.07] | 7.3 [6.89-7.7] |
| 2012 | 15.41 [14.68-16.15] | 9.52 [9.11-9.94] | 51.08 [49.62-52.55] | 7.27 [6.91-7.62] | 53.56 [51.5-55.61] | 7.04 [6.7-7.38] |
| 2013 | 15.38 [14.51-16.25] | 9.5 [8.96-10.04] | 51.03 [49.06-53] | 7.25 [6.82-7.69] | 53.42 [50.84-56] | 7 [6.57-7.44] |
| 2014 | 14.33 [13.53-15.13] | 8.81 [8.4-9.23] | 49.16 [47.62-50.68] | 6.76 [6.4-7.13] | 51.58 [49.28-53.88] | 6.54 [6.2-6.87] |
| 2016 | 10.29 [9.78-10.8] | 6.21 [5.94-6.49] | 39.55 [38.2-40.9] | 4.68 [4.45-4.91] | 41.84 [39.84-43.85] | 4.51 [4.28-4.72] |
| 2017 | 10.53 [9.99-11.06] | 6.36 [6.07-6.66] | 40.75 [39.34-42.16] | 4.91 [4.67-5.14] | 42.29 [40.22-44.37] | 4.59 [4.37-4.82] |
| 2018 | 9.81 [9.19-10.42] | 5.91 [5.58-6.24] | 38.62 [36.92-40.31] | 4.51 [4.24-4.78] | 40.25 [37.84-42.66] | 4.24 [3.99-4.44] |
| 2019 | 9.44 [8.79-10.08] | 5.68 [5.32-6.04] | 37.81 [36.06-39.56] | 4.36 [4.06-4.66] | 38.75 [36.37-41.14] | 3.99 [3.71-4.27] |

[1] MDE: AAPC = -4.9 (95% CI, -6.3 to -3.6), $p < 0.001$

[2] No MDE: AAPC = -5.2 (95% CI, -6.7 to -3.8), $p < 0.001$

[3] AUD: AAPC = -3.0 (95% CI, -3.8 to -2.1), $p < 0.001$

[4] No AUD: AAPC = -5.2 (95% CI, -6.6 to -3.7), $p < 0.001$

[5] MDE and AUD: AAPC = -3.1 (95% CI, -3.8 to -2.4), $p < 0.001$

[6] No MDE or AUD: AAPC = -5.6 (95% CI, -7.0 to -4.2), $p < 0.001$

**Supplemental Table 2.** Trends in past-year adjusted prevalence of driving under the influence of alcohol (DUIA) and 95% CI by past-year major depressive episode (MDE) and alcohol use disorder (AUD), stratified by sex, from 2005 to 2019.

**A. Male**

| Year | MDE[1] | No MDE[2] | AUD[3] | No AUD[4] | MDE and AUD[5] | No MDE or AUD[6] |
|---|---|---|---|---|---|---|
| 2005 | 24.46 [23.32-25.61] | 15.76 [15.05-16.47] | 62.77 [61.26-64.28] | 11.23 [10.65-11.81] | 67.38 [65.55-69.22] | 11.94 [11.37-12.52] |
| 2006 | 23.51 [22.36-24.65] | 15.07 [14.42-15.73] | 61.52 [59.99-63.04] | 10.71 [10.17-11.25] | 66.17 [64.11-68.23] | 11.38 [10.84-11.93] |
| 2007 | 23.72 [22.58-24.85] | 15.22 [14.57-15.87] | 61.89 [60.25-63.54] | 10.86 [10.27-11.46] | 66.29 [64.2-68.38] | 11.44 [10.87-12] |
| 2008 | 23.43 [22.28-24.57] | 15.02 [14.31-15.72] | 61.41 [59.74-63.08] | 10.67 [10.1-11.23] | 65.91 [63.79-68.03] | 11.27 [10.7-11.83] |
| 2009 | 22.49 [21.45-23.53] | 14.35 [13.72-14.98] | 60.05 [58.44-61.66] | 10.14 [9.62-10.65] | 64.64 [62.69-66.48] | 10.72 [10.2-11.23] |
| 2010 | 21.46 [20.28-22.64] | 13.63 [12.95-14.31] | 58.97 [57.25-60.69] | 9.73 [9.13-10.34] | 63.39 [61.34-65.45] | 10.21 [9.64-10.79] |
| 2011 | 21.55 [20.44-22.66] | 13.7 [13.03-14.36] | 59.71 [57.95-61.47] | 10.01 [9.43-10.58] | 63.88 [61.46-66.3] | 10.4 [9.82-10.99] |
| 2012 | 21.08 [20.1-22.06] | 13.37 [12.78-13.95] | 58.62 [57.13-60.12] | 9.61 [9.12-10.09] | 62.99 [61.02-64.96] | 10.52 [9.55-10.55] |
| 2013 | 21.04 [19.94-22.13] | 13.34 [12.62-14.05] | 58.57 [56.67-60.48] | 9.59 [9.03-10.15] | 62.86 [60.45-65.27] | 10 [9.41-10.6] |
| 2014 | 19.69 [18.63-20.76] | 12.4 [11.82-12.99] | 56.74 [55.16-58.32] | 8.96 [8.47-9.45] | 61.12 [58.9-63.34] | 9.36 [8.87-9.84] |
| 2016 | 14.39 [13.7-15.09] | 8.85 [8.46-9.24] | 47.02 [45.58-48.47] | 6.24 [5.93-6.56] | 51.5 [49.42-53.57] | 6.52 [6.21-6.82] |
| 2017 | 14.71 [13.98-15.44] | 9.06 [8.64-9.48] | 48.26 [46.76-49.77] | 6.54 [6.23-6.86] | 51.96 [49.82-54.1] | 6.63 [6.31-6.96] |
| 2018 | 13.75 [12.92-14.57] | 8.43 [7.98-8.88] | 46.05 [44.26-47.83] | 6.02 [5.67-6.37] | 49.85 [47.36-52.34] | 6.13 [5.78-6.47] |
| 2019 | 13.25 [12.37-14.13] | 8.11 [7.6-8.61] | 45.2 [43.32-47.08] | 5.83 [5.43-6.23] | 48.29 [45.77-50.81] | 5.78 [5.38-6.18] |

[1] MDE: AAPC=-4.7 (95% CI, -6.0 to -3.5), $p < 0.001$

[2] No MDE: AAPC = -5.1 (95% CI, -6.5 to -3.8), $p < 0.001$

[3] AUD: AAPC = -2.6 (95% CI, -3.3 to -1.8), $p < 0.001$

[4] No AUD: AAPC = -5.1 (95% CI, -6.6 to -3.5), $p < 0.001$

[5] MDE and AUD: AAPC = -2.5 (95% CI, -3.1 to -1.9), $p < 0.001$

[6] No MDE or AUD: AAPC = -5.5 (95% CI, -6.9 to -4.1), $p < 0.001$

**B. Female**

| Year | MDE[1] | No MDE[2] | AUD[3] | No AUD[4] | MDE and AUD[5] | No MDE or AUD[6] |
|---|---|---|---|---|---|---|
| 2005 | 13.4 [12.73-14.08] | 8.21 [7.83-8.59] | 48.33 [46.81-49.86] | 6.56 [6.22-6.9] | 49.35 [47.27-51.42] | 6.01 [5.71-6.32] |
| 2006 | 12.81 [12.12-13.49] | 7.82 [7.46-8.18] | 47 [45.43-48.57] | 6.24 [5.92-6.56] | 47.98 [45.71-50.26] | 5.71 [5.43-5.99] |
| 2007 | 12.94 [12.21-13.66] | 7.9 [7.51-8.29] | 47.4 [45.65-49.15] | 6.33 [5.96-6.7] | 48.12 [45.73-40.41] | 5.74 [5.42-6.06] |
| 2008 | 12.76 [12.02-13.49] | 7.79 [7.37-8.2] | 46.89 [45.12-48.66] | 6.21 [5.86-6.57] | 47.7 [45.3-50.09] | 5.65 [5.33-5.97] |
| 2009 | 12.18 [11.54-12.82] | 7.41 [7.06-7.77] | 45.48 [43.81-47.14] | 5.89 [5.56-6.21] | 46.29 [44.13-48.45] | 5.36 [5.07-5.64] |
| 2010 | 11.55 [10.91-12.19] | 7.01 [6.69-7.33] | 44.37 [42.8-45.93] | 5.65 [5.32-5.97] | 44.96 [42.91-47] | 5.09 [4.83-5.35] |
| 2011 | 11.61 [10.95-12.26] | 7.05 [6.69-7.4] | 45.12 [43.37-46.87] | 5.81 [5.47-6.15] | 45.48 [42.91-48.05] | 5.19 [4.9-5.49] |
| 2012 | 11.32 [10.74-11.9] | 6.87 [6.55-7.19] | 44.01 [42.53-45.5] | 5.57 [5.28-5.86] | 44.53 [42.46-46.6] | 5.01 [4.75-5.26] |
| 2013 | 11.29 [10.59-12] | 6.85 [6.42-7.28] | 43.96 [41.94-45.98] | 5.56 [5.19-5.92] | 44.39 [41.76-47.02] | 4.98 [4.64-5.32] |
| 2014 | 10.49 [9.87-11.11] | 6.34 [6.02-6.66] | 42.12 [40.6-43.64] | 5.18 [4.89-5.47] | 42.58 [40.3-44.85] | 4.64 [4.39-4.89] |
| 2016 | 7.44 [7.04-7.83] | 4.44 [4.22-4.65] | 33 [31.69-34.31] | 3.56 [3.37-3.76] | 33.36 [31.49-35.24] | 3.18 [3.02-3.35] |
| 2017 | 7.61 [7.2-8.03] | 4.54 [4.31-4.77] | 34.11 [32.74-35.47] | 3.74 [3.55-3.93] | 33.78 [31.83-35.73] | 3.24 [3.07-3.41] |
| 2018 | 7.08 [6.6-7.55] | 4.21 [3.96-4.47] | 32.14 [30.51-33.76] | 3.43 [3.21-3.65] | 31.92 [29.69-34.15] | 2.99 [2.8-3.18] |
| 2019 | 6.8 [6.31-7.29] | 4.04 [3.78-4.31] | 31.4 [29.75-33.04] | 3.32 [3.08-3.56] | 30.58 [28.41-32.74] | 2.81 [2.6-3.02] |

[1] MDE: AAPC = -5.2 (95% CI, -6.6 to -3.9), $p < 0.001$

[2] No MDE: AAPC = -5.4 (95% CI, -6.8 to -4.1), $p < 0.001$

[3] AUD: AAPC = -3.3 (95% CI, -4.3 to -2.4), $p < 0.001$

[4] No AUD: AAPC = -5.2 (95% CI, -6.7 to -3.7), $p < 0.001$

[5] MDE and AUD: AAPC = -3.6 (95% CI, -4.4 to -2.7), $p < 0.001$

[6] No MDE or AUD: AAPC = -5.7 (95% CI, -7.1 to -4.2), $p < 0.001$

**Supplemental Table 3.** Trends in past-year adjusted prevalence of driving under the influence of alcohol (DUIA) and 95% CI by past-year major depressive episode (MDE) and alcohol use disorder (AUD), stratified by age, from 2005 to 2019.

**A. 18 to 25 years**

| Year | MDE[1] | No MDE[2] | AUD[3] | No AUD[4] | MDE and AUD[5] | No MDE or AUD[6] |
|---|---|---|---|---|---|---|
| 2005 | 27.45 [26.19-28.72] | 17.94 [17.13-18.75] | 64.96 [63.51-66.4] | 12.21 [11.57-12.86] | 68.56 [66.64-70.47] | 12.52 [11.9-13.15] |
| 2006 | 26.42 [25.17-27.67] | 17.18 [16.44-17.92] | 63.74 [62.26-65.22] | 11.65 [11.05-12.25] | 67.37 [65.25-69.49] | 11.94 [11.35-12.53] |
| 2007 | 26.64 [25.29-28] | 17.34 [16.51-18.17] | 64.1 [62.04-65.81] | 11.82 [11.1-12.53] | 67.49 [65.25-69.73] | 11.99 [11.31-12.67] |
| 2008 | 26.33 [24.99-27.67] | 17.11 [16.25-17.97] | 63.63 [61.91-65.35] | 11.6 [10.92-12.29] | 67.11 [64.88-69.35] | 11.82 [11.16-12.47] |
| 2009 | 25.31 [24.14-26.49] | 16.37 [15.64-17.1] | 62.31 [60.72-63.9] | 11.04 [10.45-11.62] | 65.86 [63.81-67.91] | 11.24 [10.66-11.82] |
| 2010 | 24.2 [22.95-25.45] | 15.57 [14.85-16.29] | 61.25 [59.59-62.9] | 10.6 [9.94-11.26] | 64.64 [62.53-66.75] | 10.72 [10.11-11.33] |
| 2011 | 24.3 [23.04-25.56] | 15.64 [14.87-16.41] | 61.97 [60.25-63.69] | 10.89 [10.25-11.54] | 65.12 [62.64-67.6] | 10.92 [10.87-11.55] |
| 2012 | 23.79 [22.69-24.88] | 15.27 [14.6-15.94] | 60.91 [59.49-62.32] | 10.47 [9.94-11] | 64.24 [62.21-66.27] | 10.55 [10.03-11.08] |
| 2013 | 23.74 [22.4-25.08] | 15.24 [14.34-16.14] | 60.86 [58.96-62.76] | 10.45 [9.81-11.09] | 64.11 [61.6-66.63] | 10.5 [9.83-11.17] |
| 2014 | 22.27 [21.08-23.45] | 14.2 [13.53-14.87] | 59.05 [57.5-60.61] | 9.77 [9.21-10.32] | 62.4 [60.09-64.7] | 9.83 [9.29-10.36] |
| 2016 | 16.42 [15.59-17.25] | 10.19 [9.71-10.67] | 49.39 [47.85-50.93] | 6.82 [6.44-7.21] | 52.84 [50.61-55.08] | 6.85 [6.5-7.21] |
| 2017 | 16.77 [15.9-17.64] | 10.42 [9.91-10.94] | 50.64 [49.05-52.22] | 7.15 [6.76-7.53] | 53.31 [50.99-55.62] | 6.97 [6.59-7.35] |
| 2018 | 15.7 [14.73-16.68] | 9.71 [9.17-10.26] | 48.42 [46.55-50.28] | 6.58 [6.16-7] | 51.2 [48.56-53.85] | 6.45 [6.05-6.84] |
| 2019 | 15.14 [14.16-16.12] | 9.34 [8.77-9.92] | 47.56 [45.7-49.43] | 6.37 [5.93-6.81] | 49.64 [47.04-52.24] | 6.08 [5.66-6.5] |

[1] MDE: AAPC = -4.6 (95% CI, -5.7 to -3.4), $p < 0.001$

[2] No MDE: AAPC = -5.0 (95% CI, -6.3 to -3.7), $p < 0.001$

[3] AUD: AAPC = -2.4 (95% CI, -3.1 to -1.7), $p < 0.001$

[4] No AUD: AAPC = -4.9 (95% CI, -6.3 to -3.5), $p < 0.001$

[5] MDE and AUD: AAPC = -2.4 (95% CI, -3.0 to -1.9), $p < 0.001$

[6] No MDE or AUD: AAPC = -5.4 (95% CI, -6.7 to -4.1), $p < 0.001$

**B. 26 to 34 years**

| Year | MDE[1] | No MDE[2] | AUD[3] | No AUD[4] | MDE and AUD[5] | No MDE or AUD[6] |
|---|---|---|---|---|---|---|
| 2005 | 26.88 [25.72-28.04] | 17.52 [16.73-18.3] | 65.7 [64.2-67.2] | 12.57 [11.93-13.2] | 68.55 [66.77-70.33] | 12.52 [11.89-13.15] |
| 2006 | 25.86 [24.69-27.04] | 16.77 [16.04-17.51] | 64.49 [62.96-66.01] | 11.99 [11.4-12.58] | 67.36 [65.37-69.35] | 11.93 [11.35-12.52] |
| 2007 | 26.09 [24.89-27.28] | 16.93 [16.18-17.69] | 64.85 [63.19-66.51] | 12.16 [11.49-12.83] | 67.48 [65.44-69.51] | 11.99 [11.38-12.6] |
| 2008 | 25.78 [24.52-27.03] | 16.71 [15.86-17.55] | 64.38 [62.65-66.12] | 11.94 [11.28-12.61] | 67.11 [65-69.21] | 11.81 [11.17-12.46] |
| 2009 | 24.77 [23.62-25.93] | 15.98 [15.21-16.75] | 63.07 [61.31-64.83] | 11.36 [10.72-12] | 65.85 [63.85-67.85] | 11.24 [10.61-11.87] |
| 2010 | 23.67 [22.46-24.89] | 15.19 [14.45-15.94] | 62.02 [60.28-63.75] | 10.91 [10.24-11.59] | 64.63 [62.64-66.61] | 10.71 [10.1-11.33] |
| 2011 | 23.77 [22.56-24.99] | 15.26 [14.47-16.06] | 62.74 [60.86-64.61] | 11.22 [10.52-11.91] | 65.11 [62.69-67.53] | 10.92 [10.25-11.58] |
| 2012 | 23.27 [22.29-24.24] | 14.9 [14.27-15.53] | 61.68 [60.2-63.16] | 10.78 [10.26-11.3] | 64.23 [62.36-66.1] | 10.55 [10.03-11.06] |
| 2013 | 23.22 [21.98-24.46] | 14.87 [14-15.74] | 61.63 [59.57-63.69] | 10.76 [10.06-11.45] | 64.1 [61.66-66.55] | 10.5 [9.8-11.19] |
| 2014 | 21.77 [20.66-22.89] | 13.85 [13.19-14.51] | 59.84 [58.21-61.48] | 10.06 [9.5-10.62] | 62.39 [60.23-64.54] | 9.82 [9.3-10.34] |
| 2016 | 16.03 [15.28-16.78] | 9.93 [9.47-10.39] | 50.21 [48.63-51.79] | 7.03 [6.66-7.4] | 52.83 [50.77-54.89] | 6.85 [6.51-7.2] |
| 2017 | 16.37 [15.6-17.15] | 10.16 [9.67-10.64] | 51.45 [49.86-53.05] | 7.37 [7.01-7.73] | 53.3 [51.2-55.4] | 6.97 [6.62-7.32] |
| 2018 | 15.32 [14.45-16.19] | 9.46 [8.96-9.97] | 49.23 [47.35-51.11] | 6.78 [6.38-7.19] | 51.19 [48.74-53.65] | 6.44 [6.07-6.82] |
| 2019 | 14.77 [13.86-15.69] | 9.1 [8.54-9.66] | 48.37 [46.47-50.28] | 6.57 [6.13-7.01] | 49.63 [47.17-52.08] | 6.08 [5.66-6.49] |

[1] MDE: AAPC = -4.6 (95% CI, -5.8 to -3.4), $p < 0.001$

[2] No MDE: AAPC = -5.0 (95% CI, -6.3 to -3.7), $p < 0.001$

[3] AUD: AAPC = -2.3 (95% CI, -3.0 to -1.7), $p < 0.001$

[4] No AUD: AAPC = -4.9 (95% CI, -6.3 to -3.5), $p < 0.001$

[5] MDE and AUD: AAPC = -2.4 (95% CI, -3.0 to -1.9), $p < 0.001$

[6] No MDE or AUD: AAPC = -5.4 (95% CI, -6.7 to -4.0), $p < 0.001$

**C. 35 to 49 years**

| Year | MDE[1] | No MDE[2] | AUD[3] | No AUD[4] | MDE and AUD[5] | No MDE or AUD[6] |
|---|---|---|---|---|---|---|
| 2005 | 21.02 [20.01-22.04] | 13.33 [12.7-13.95] | 59.66 [58.05-61.27] | 9.99 [9.44-10.54] | 62.03 [60.08-63.98] | 9.69 [9.19-10.19] |
| 2006 | 20.17 [19.18-21.15] | 12.73 [12.18-13.29] | 58.37 [56.82-59.92] | 9.52 [9.05-10] | 60.73 [58.63-62.84] | 9.22 [8.79-9.65] |
| 2007 | 20.35 [19.28-21.42] | 12.86 [12.23-13.49] | 58.76 [56.97-60.55] | 9.66 [9.08-10.23] | 60.86 [58.6-63.12] | 9.26 [8.75-9.78] |
| 2008 | 20.09 [19.08-21.1] | 12.68 [12.07-13.29] | 58.26 [56.57-59.96] | 9.48 [8.98-9.98] | 60.46 [58.27-62.65] | 9.12 [8.66-9.59] |
| 2009 | 19.25 [18.37-20.13] | 12.1 [11.58-12.63] | 56.88 [55.29-58.46] | 9.01 [8.57-9.44] | 59.1 [57.14-61.06] | 8.67 [8.26-9.07] |
| 2010 | 18.34 [17.4-19.28] | 11.48 [10.98-11.99] | 55.77 [54.16-57.38] | 8.64 [8.15-9.14] | 57.79 [55.82-59.77] | 8.25 [7.83-8.67] |
| 2011 | 18.42 [17.46-19.38] | 11.54 [10.97-12.1] | 56.52 [54.76-58.28] | 8.89 [8.39-9.39] | 58.31 [55.83-60.78] | 8.41 [7.95-8.89] |
| 2012 | 18 [17.15-18.86] | 11.25 [10.75-11.76] | 55.42 [53.89-56.94] | 8.53 [8.09-8.97] | 57.37 [55.36-59.38] | 8.12 [7.71-8.53] |
| 2013 | 17.96 [16.98-18.95] | 11.23 [10.59-11.87] | 55.36 [53.38-57.34] | 8.51 [8-9.03] | 57.24 [54.73-59.74] | 8.08 [7.57-8.58] |
| 2014 | 16.77 [15.84-17.71] | 10.43 [9.92-10.94] | 53.5 [51.89-55.12] | 7.95 [7.5-8.4] | 55.42 [53.16-57.68] | 7.55 [7.14-7.95] |
| 2016 | 12.14 [11.55-12.74] | 7.39 [7.06-7.73] | 43.78 [42.33-45.23] | 5.52 [5.24-5.8] | 45.64 [43.63-47.64] | 5.22 [4.97-5.48] |
| 2017 | 12.41 [11.79-13.04] | 7.57 [7.21-7.93] | 45.01 [43.47-46.55] | 5.79 [5.49-6.08] | 46.1 [44-48.19] | 5.32 [5.05-5.59] |
| 2018 | 11.58 [10.85-12.32] | 7.03 [6.63-7.44] | 42.82 [40.97-44.66] | 5.32 [4.99-5.65] | 44.01 [41.54-46.48] | 4.91 [4.61-5.21] |
| 2019 | 11.15 [10.41-11.9] | 6.76 [6.33-7.19] | 41.98 [40.13-43.83] | 5.15 [4.79-5.51] | 42.48 [40.06-44.89] | 4.62 [4.3-4.94] |

[1] MDE: AAPC = -4.9 (95% CI, -6.2 to -3.6), $p < 0.001$

[2] No MDE: AAPC= -5.2 (95% CI, -3.8 to -7.3), $p < 0.001$

[3] AUD: AAPC = -2.7 (95% CI, -3.5 to -1.9), $p < 0.001$

[4] No AUD: AAPC = -5.0 (95% CI, -6.4 to -3.6), $p < 0.001$

[5] MDE and AUD: AAPC = -2.8 (95% CI, -3.5 to -2.2), $p < 0.001$

[6] No MDE or AUD: AAPC = -5.5 (95% CI, -6.9 to -4.1), $p < 0.001$

**D. ≥ 50 years**

| Year | MDE[1] | No MDE[2] | AUD[3] | No AUD[4] | MDE and AUD[5] | No MDE or AUD[6] |
|---|---|---|---|---|---|---|
| 2005 | 12 [11.33-12.66] | 7.3 [6.93-7.67] | 45.41 [43.81-47.01] | 5.88 [5.55-6.2] | 48.23 [46.09-50.37] | 5.76 [5.46-6.06] |
| 2006 | 11.45 [10.77-12.14] | 6.95 [6.6-7.31] | 44.09 [42.41-45.78] | 5.59 [5.26-5.91] | 46.87 [44.48-49.25] | 5.47 [5.17-5.78] |
| 2007 | 11.57 [10.9-12.24] | 7.03 [6.68-7.37] | 44.49 [42.75-46.23] | 5.67 [5.33-6.01] | 47 [44.6-49.4] | 5.5 [5.2-5.81] |
| 2008 | 11.41 [10.71-12.11] | 6.92 [6.53-7.31] | 43.98 [42.16-45.8] | 5.56 [5.22-5.9] | 46.58 [44.12-49.04] | 5.42 [5.1-5.73] |
| 2009 | 10.88 [10.25-11.51] | 6.59 [6.24-6.93] | 42.59 [40.85-44.32] | 5.27 [4.96-5.58] | 45.18 [42.95-47.41] | 5.13 [4.84-5.43] |
| 2010 | 10.31 [9.65-10.98] | 6.23 [5.89-6.57] | 41.49 [39.77-43.21] | 5.05 [4.72-5.39] | 43.85 [41.65-46.05] | 4.88 [4.59-5.17] |
| 2011 | 10.36 [9.74-10.99] | 6.26 [5.93-6.59] | 42.24 [40.43-44.04] | 5.2 [4.88-5.53] | 44.37 [41.74-46.99] | 4.98 [4.68-5.27] |
| 2012 | 10.11 [9.52-10.69] | 6.1 [5.78-6.41] | 41.14 [39.54-42.75] | 4.98 [4.59-5.28] | 43.42 [41.24-45.6] | 4.8 [4.52-5.07] |
| 2013 | 10.08 [9.44-10.73] | 6.08 [5.7-6.46] | 41.09 [39.08-43.1] | 4.97 [4.64-5.31] | 43.29 [40.65-45.93] | 4.77 [4.45-5.1] |
| 2014 | 9.35 [8.75-9.96] | 5.62 [5.32-5.93] | 39.29 [37.71-40.87] | 4.63 [4.35-4.91] | 41.48 [39.14-43.83] | 4.44 [4.19-4.71] |
| 2016 | 6.61 [6.23-6.99] | 3.93 [3.72-4.13] | 30.46 [29.14-31.78] | 3.18 [3-3.37] | 32.38 [30.45-34.3] | 3.05 [2.88-3.21] |
| 2017 | 6.77 [6.36-7.17] | 4.02 [3.81-4.24] | 31.52 [30.15-32.9] | 3.34 [3.16-3.52] | 32.78 [30.8-34.77] | 3.1 [2.93-3.28] |
| 2018 | 6.29 [5.84-6.73] | 3.73 [3.5-3.96] | 29.63 [28.05-31.22] | 3.06 [2.86-3.27] | 30.95 [28.71-33.19] | 2.86 [2.67-3.04] |
| 2019 | 6.04 [5.57-6.51] | 3.58 [3.32-3.84] | 28.93 [27.25-30.6] | 2.96 [2.73-3.19] | 29.63 [27.4-31.86] | 2.69 [2.48-2.9] |

[1] MDE: AAPC = -5.3 (95% CI, -6.7 to -3.9), $p < 0.001$

[2] No MDE: AAPC = -5.5 (95% CI, -6.9 to -4.1), $p < 0.001$

[3] AUD: AAPC = -3.4 (95% CI, -4.4 to -2.4), $p < 0.001$

[4] No AUD: AAPC = -5.2 (95% CI, -6.6 to -3.7), $p < 0.001$

[5] MDE and AUD: AAPC = -3.6 (95% CI, -4.5 to -2.8), $p < 0.001$

[6] No MDE or AUD: AAPC = -5.7 (95% CI, -7.2 to -4.3), $p < 0.001$

**Supplemental Table 4.** Trends in past-year adjusted prevalence of driving under the influence of alcohol (DUIA) and 95% CI by past-year major depressive episode (MDE) and alcohol use disorder (AUD), stratified by race and ethnicity, from 2005 to 2019.

**A. Non-Hispanic Whites**

| Year | MDE[1] | No MDE[2] | AUD[3] | No AUD[4] | MDE and AUD[5] | No MDE or AUD[6] |
|---|---|---|---|---|---|---|
| 2005 | 21.22 [20.21-22.22] | 13.46 [12.84-14.99] | 60.13 [58.6-61.67] | 10.17 [9.64-10.69] | 62.51 [60.59-64.43] | 9.87 [9.37-10.36] |
| 2006 | 20.36 [19.4-21.31] | 12.86 [12.33-13.4] | 58.85 [57.35-60.35] | 9.69 [9.22-10.16] | 61.22 [59.14-63.3] | 9.39 [8.96-0.92] |
| 2007 | 20.54 [19.56-21.52] | 12.99 [12.43-13.55] | 59.23 [57.62-60.85] | 9.83 [9.31-10.35] | 61.35 [59.2-63.49] | 9.44 [8.98-9.9] |
| 2008 | 20.28 [19.25-21.31] | 12.81 [12.18-13.44] | 58.74 [57.02-60.45] | 9.65 [9.13-10.17] | 60.94 [58.73-63.16] | 9.29 [8.81-9.78] |
| 2009 | 19.44 [18.53-20.34] | 12.23 [11.68-12.78] | 56.36 [55.72-58.99] | 9.17 [8.7-9.63] | 59.6 [57.59-61.6] | 8.83 [8.39-9.27] |
| 2010 | 18.52 [17.55-19.48] | 11.6 [11.07-12.13] | 56.25 [54.63-57.88] | 8.8 [8.29-9.31] | 58.29 [56.29-60.29] | 8.41 [7.96-8.85] |
| 2011 | 18.6 [17.69-19.51] | 11.66 [11.13-12.19] | 57 [55.31-58.7] | 9.05 [8.56-9.53] | 58.8 [56.38-61.22] | 8.57 [8.12-9.02] |
| 2012 | 18.18 [17.34-19.01] | 11.37 [10.88-11.87] | 55.9 [54.42-57.38] | 8.68 [8.25-9.11] | 57.87 [55.9-69.84] | 8.27 [7.87-8.68] |
| 2013 | 18.14 [17.14-19.13] | 11.34 [10.69-12] | 55.85 [53.84-57.86] | 8.67 [8.13-9.21] | 57.73 [55.21-60.26] | 8.23 [7.71-8.76] |
| 2014 | 16.94 [16.02-17.84] | 10.54 [10.04-11.03] | 53.99 [52.43-55.55] | 8.09 [7.65-8.53] | 55.92 [53.7-58.15] | 7.69 [7.29-8.09] |
| 2016 | 12.27 [11.68-12.85] | 7.47 [7.14-7.81] | 44.26 [42.83-45.69] | 5.62 [5.34-5.91] | 46.14 [44.14-48.15] | 5.33 [5.07-5.58] |
| 2017 | 12.54 [11.94-13.14] | 7.65 [7.31-7.99] | 45.49 [44.06-46.93] | 5.89 [5.62-6.16] | 46.61 [44.57-48.64] | 5.42 [5.17-5.68] |
| 2018 | 11.7 [10.99-12.41] | 7.11 [6.72-7.5] | 43.3 [41.54-45.06] | 5.42 [5.1-5.74] | 44.51 [42.09-46.94] | 5 [5.71-5.29] |
| 2019 | 11.27 [10.52-12.01] | 6.83 [6.4-7.26] | 42.46 [40.63-44.29] | 5.26 [4.89-5.6] | 42.97 [40.56-45.39] | 4.72 [4.39-5.04] |

[1] MDE: AAPC = -4.9 (95% CI, -6.1 to -3.6), $p < 0.001$

[2] No MDE: AAPC = -5.2 (95% CI, -6.5 to -3.9), $p < 0.001$

[3] AUD: AAPC = -2.7 (95% CI, -3.5 to -1.9), $p < 0.001$

[4] No AUD: AAPC = -5.1 (95% CI, -6.6 to -3.6), $p < 0.001$

[5] MDE and AUD: AAPC = -2.8 (95% CI, -3.5 to -2.2), $p < 0.001$

[6] No MDE or AUD: AAPC = -5.5 (95% CI, -6.9 to -4.2), $p < 0.001$

**B. Non-Hispanic Black**

| Year | MDE[1] | No MDE[2] | AUD[3] | No AUD[4] | MDE and AUD[5] | No MDE or AUD[6] |
|---|---|---|---|---|---|---|
| 2005 | 13.35 [12.53-14.16] | 8.17 [7.71-8.63] | 47.31 [45.46-49.16] | 6.31 [5.89-6.73] | 51.33 [48.97-53.69] | 6.48 [6.08-6.87] |
| 2006 | 12.75 [11.86-13.64] | 7.78 [7.29-8.28] | 45.98 [43.97-47.99] | 6 [5.57-6.44] | 49.96 [47.26-52.66] | 6.15 [5.73-6.58] |
| 2007 | 12.88 [12.01-13.75] | 7.87 [7.39-8.35] | 46.38 [44.26-48.5] | 6.09 [5.63-6.55] | 50.1 [47.36-52.83] | 6.18 [5.75-6.62] |
| 2008 | 12.7 [11.85-13.55] | 7.75 [7.27-8.23] | 45.87 [43.9-47.84] | 5.98 [5.57-6.38] | 49.67 [47.04-52.31] | 6.09 [5.69-6.48] |
| 2009 | 12.12 [11.35-12.89] | 7.38 [6.95-7.81] | 44.46 [42.52-46.4] | 5.67 [5.28-6.05] | 48.27 [45.81-50.72] | 5.77 [5.4-6.14] |
| 2010 | 11.5 [10.66-12.34] | 6.98 [6.52-7.44] | 43.36 [41.4-45.32] | 5.43 [5.02-5.84] | 46.92 [44.41-49.43] | 5.49 [5.1-5.87] |
| 2011 | 11.55 [10.76-12.35] | 7.02 [6.58-7.46] | 44.11 [42.01-46.21] | 5.59 [5.17-6.01] | 47.45 [44.54-40.36] | 5.6 [5.19-6] |
| 2012 | 11.27 [10.54-11.99] | 6.83 [6.43-7.24] | 43 [41.22-44.79] | 5.36 [5-5.71] | 46.49 [44.06-48.92] | 5.4 [5.04-5.75] |
| 2013 | 11.24 [10.49-12] | 6.82 [6.37-7.27] | 42.95 [40.84-45.07] | 5.35 [4.97-5.73] | 46.35 [43.56-49.15] | 5.37 [4.99-5.75] |
| 2014 | 10.44 [9.7-11.18] | 6.31 [5.92-6.7] | 41.13 [39.32-42.93] | 4.98 [4.63-5.33] | 44.52 [41.92-47.12] | 5.01 [4.67-5.34] |
| 2016 | 7.4 [6.92-7.89] | 4.41 [4.15-4.68] | 32.1 [30.52-33.68] | 3.43 [3.19-3.66] | 35.15 [32.92-37.38] | 3.44 [3.21-3.66] |
| 2017 | 7.58 [7.04-8.12] | 4.52 [4.22-4.82] | 33.19 [31.46-34.92] | 3.59 [3.34-3.85] | 35.57 [33.2-37.95] | 3.5 [3.25-3.75] |
| 2018 | 7.04 [6.51-7.58] | 4.19 [3.91-4.48] | 31.25 [29.46-33.04] | 3.3 [3.05-3.54] | 33.66 [31.17-36.16] | 3.22 [2.99-3.46] |
| 2019 | 6.77 [6.2-7.34] | 4.03 [3.71-4.34] | 30.52 [28.6-32.44] | 3.19 [2.91-3.47] | 32.28 [29.75-34.82] | 3.04 [2.77-3.3] |

[1] MDE: AAPC = -5.2 (95% CI, -6.5 to -3.9), $p < 0.001$

[2] No MDE: AAPC = -5.4 (95% CI, -6.8 to -4.0), $p < 0.001$

[3] AUD: AAPC = -3.4 (95% CI, -4.3 to -2.4), $p < 0.001$

[4] No AUD: AAPC = -5.2 (95% CI, -6.7 to -3.7), $p < 0.001$

[5] MDE and AUD: AAPC = -3.5 (95% CI, -4.3 to -2.7), $p < 0.001$

[6] No MDE or AUD: AAPC = -5.7 (95% CI, -7.0 to -4.3), $p < 0.001$

**C. Hispanic**

| Year | MDE[1] | No MDE[2] | AUD[3] | No AUD[4] | MDE and AUD[5] | No MDE or AUD[6] |
|---|---|---|---|---|---|---|
| 2005 | 14.41 [13.6-15.22] | 8.86 [8.4-9.32] | 48.05 [46.33-49.79] | 6.49 [6.13-6.86] | 52.17 [49.86-54.49] | 6.68 [6.32-7.05] |
| 2006 | 13.77 [12.91-14.64] | 8.45 [7.97-8.92] | 46.73 [44.85-48.62] | 6.18 [5.8-6.56] | 50.81 [48.2-53.41] | 6.35 [5.97-6.73] |
| 2007 | 13.91 [12.99-14.83] | 8.54 [8.02-9.05] | 47.13 [44.98-49.27] | 6.27 [5.82-6.72] | 50.94 [48.2-53.68] | 6.38 [5.95-6.81] |
| 2008 | 13.72 [12.85-14.59] | 8.41 [7.91-8.91] | 46.61 [44.6-48.63] | 6.15 [5.75-6.55] | 50.52 [47.85-53.18] | 6.28 [5.89-6.67] |
| 2009 | 13.1 [12.29-13.91] | 8.01 [7.55-8.47] | 45.21 [43.23-47.18] | 5.83 [5.46-6.2] | 49.11 [46.61-51.61] | 5.96 [5.58-6.33] |
| 2010 | 12.44 [11.49-13.28] | 7.58 [7.13-8.04] | 44.1 [42.04-46.15] | 5.59 [5.17-6] | 47.76 [45.25-50.27] | 5.66 [5.28-6.04] |
| 2011 | 12.5 [11.59-13.4] | 7.62 [7.1-8.14] | 44.85 [42.63-47.07] | 5.75 [5.32-6.18] | 48.29 [45.29-51.29] | 5.78 [5.35-6.2] |
| 2012 | 12.19 [11.45-12.93] | 7.42 [7-7.84] | 43.74 [41.93-45.55] | 5.55 [5.17-5.85] | 47.33 [44.91-49.75] | 5.57 [5.23-5.91] |
| 2013 | 12.16 [11.31-13.01] | 7.41 [6.89-7.92] | 43.69 [41.48-45.9] | 5.5 [5.11-5.89] | 47.19 [44.31-50.07] | 5.54 [5.14-5.94] |
| 2014 | 11.3 [10.55-12.06] | 6.86 [6.46-7.26] | 41.85 [39.99-43.72] | 5.12 [4.78-5.47] | 45.36 [42.75-47.96] | 5.17 [4.84-5.49] |
| 2016 | 8.04 [7.54-8.54] | 4.81 [4.53-5.08] | 32.76 [31.22-34.3] | 3.53 [3.31-3.74] | 35.92 [33.73-38.11] | 3.55 [3.35-3.75] |
| 2017 | 8.23 [7.67-8.79] | 4.92 [4.61-5.24] | 33.86 [32.12-35.61] | 3.7 [3.46-3.94] | 36.36 [33.99-38.71] | 3.61 [3.38-3.85] |
| 2018 | 7.65 [7.06-8.24] | 4.57 [0-4.89] | 31.9 [39.98-33.82] | 3.4 [3.14-3.65] | 34.42 [31.82-37.02] | 3.33 [3.09-3.58] |
| 2019 | 7.35 [0.68-7.93] | 4.38 [4.07-4.7] | 31.16 [29.32-33] | 3.28 [3.03-3.54] | 33.02 [30.55-35.49] | 3.14 [2.89-3.38] |

[1] MDE: AAPC = -5.2 (95% CI, -6.5 to -3.8), $p < 0.001$

[2] No MDE: AAPC = -5.4 (95% CI, -6.7 to -4.0), $p < 0.001$

[3] AUD: AAPC = -3.4 (95% CI, -4.3 to -2.4), $p < 0.001$

[4] No AUD: AAPC = -5.2 (95% CI, -6.7 to -3.7), $p < 0.001$

[5] MDE and AUD: AAPC = -3.4 (95% CI, -4.2 to -2.6), $p < 0.001$

[6] No MDE or AUD: AAPC = -5.7 (95% CI, -7.1 to -4.2), $p < 0.001$

**D. Other Race**

| Year | MDE[1] | No MDE[2] | AUD[3] | No AUD[4] | MDE and AUD[5] | No MDE or AUD[6] |
|---|---|---|---|---|---|---|
| 2005 | 9.84 [9.09-10.59] | 5.93 [5.51-6.35] | 38.7 [36.69-40.7] | 4.52 [4.15-4.89] | 41.94 [39.47-44.41] | 4.53 [4.19-4.87] |
| 2006 | 9.39 [8.62-10.16] | 5.65 [5.22-6.07] | 37.44 [35.4-39.48] | 4.3 [3.94-4.66] | 40.62 [37.92-43.32] | 4.3 [3.96-4.64] |
| 2007 | 9.49 [8.7-10.27] | 5.71 [5.27-6.14] | 37.81 [35.63-40] | 4.36 [3.98-4.75] | 40.75 [37.96-43.53] | 4.32 [3.96-4.68] |
| 2008 | 9.35 [8.58-10.12] | 5.62 [5.19-6.06] | 37.33 [35.14-39.52] | 4.28 [3.9-4.65] | 40.34 [37.57-43.11] | 4.25 [3.9-4.6] |
| 2009 | 8.91 [8.22-9.59] | 5.35 [4.96-5.73] | 36.01 [34.02-38.01] | 4.05 [3.72-4.38] | 38.99 [36.49-41.49] | 4.03 [3.72-4.34] |
| 2010 | 8.43 [7.76-9.1] | 5.05 [4.7-5.41] | 34.98 [33.1-36.87] | 3.88 [3.56-4.2] | 37.72 [35.32-40.11] | 3.82 [3.53-4.12] |
| 2011 | 8.48 [7.78-9.17] | 5.08 [4.69-5.46] | 35.68 [33.55-37.82] | 4 [3.65-4.35] | 38.21 [35.32-41.1] | 3.9 [3.57-4.34] |
| 2012 | 8.26 [7.6-8.92] | 4.94 [4.58-5.31] | 34.66 [32.7-36.62] | 3.83 [3.5-4.15] | 37.31 [34.84-39.78] | 3.76 [3.46-4.06] |
| 2013 | 8.24 [7.56-8.92] | 4.93 [4.54-5.33] | 34.61 [32.47-36.75] | 3.82 [3.49-4.15] | 37.18 [34.37-39.99] | 3.74 [3.42-4.06] |
| 2014 | 7.63 [6.97-8.29] | 4.56 [4.2-4.91] | 32.93 [31.01-34.85] | 3.55 [3.25-3.86] | 35.47 [32.88-38.07] | 3.48 [3.2-3.77] |
| 2016 | 5.36 [4.92-5.81] | 3.17 [2.93-3.41] | 24.94 [23.34-26.55] | 2.43 [2.22-2.64] | 27.07 [24.95-29.2] | 2.38 [2.19-2.57] |
| 2017 | 5.49 [5.05-5.94] | 3.25 [3-3.49] | 25.89 [24.29-27.48] | 2.55 [2.35-2.76] | 27.44 [25.29-29.6] | 2.42 [2.23-2.62] |
| 2018 | 5.1 [4.64-5.56] | 3.01 [2.77-3.25] | 24.22 [22.46-25.98] | 2.34 [2.13-2.56] | 25.79 [23.5-28.09] | 2.23 [2.04-2.42] |
| 2019 | 4.9 [4.43-5.36] | 2.89 [2.63-3.14] | 23.59 [21.83-25.36] | 2.26 [2.04-2.49] | 24.62 [22.38-26.85] | 2.1 [1.9-2.3] |

[1] MDE: AAPC = -5.3 (95% CI, -6.6 to -4.0), $p < 0.001$

[2] No MDE: AAPC = -5.5 (95% CI, -6.8 to -4.1), $p < 0.001$

[3] AUD: AAPC = -3.8 (95% CI, -4.8 to -2.7), $p < 0.001$

[4] No AUD: AAPC = -5.3 (95% CI, -6.7 to -3.8), $p < 0.001$

[5] MDE and AUD: AAPC = -4.0 (95% CI, -4.9 to -3.0), $p < 0.001$

[6] No MDE or AUD: AAPC = -5.7 (95% CI, -7.0 to -4.3), $p < 0.001$

**Supplemental Table 5.** Trends in past-year adjusted prevalence of driving under the influence of alcohol (DUIA) and 95% CI by past-year major depressive episode (MDE) and alcohol use disorder (AUD), stratified by marital status, from 2005 to 2019.

**A. Never married**

| Year | MDE[1] | No MDE[2] | AUD[3] | No AUD[4] | MDE and AUD[5] | No MDE or AUD[6] |
|---|---|---|---|---|---|---|
| 2005 | 22.81 [21.71-23.91] | 14.58 [13.89-15.27] | 60.24 [58.62-61.86] | 10.21 [9.62-10.8] | 63.32 [61.35-65.29] | 10.18 [9.62-10.74] |
| 2006 | 21.9 [20.8-23] | 13.94 [13.3-14.58] | 58.96 [57.37-60.54] | 9.73 [9.2-10.26] | 62.04 [59.9-64.19] | 9.69 [9.19-10.19] |
| 2007 | 22.1 [20.96-23.24] | 14.08 [13.4-14.75] | 59.34 [57.61-61.07] | 9.87 [9.28-10.46] | 62.17 [59.95-64.4] | 9.74 [9.2-10.28] |
| 2008 | 21.82 [20.7-22.94] | 13.88 [13.19-14.58] | 58.85 [57.1-60.6] | 9.69 [9.13-10.25] | 61.77 [59.53-64.1] | 9.59 [9.06-10.13] |
| 2009 | 20.93 [19.91-21.94] | 13.26 [12.64-13.88] | 57.47 [55.76-59.18] | 9.2 [8.69-9.72] | 60.43 [58.38-62.49] | 9.12 [8.62-9.61] |
| 2010 | 19.95 [18.87-21.04] | 12.59 [11.97-13.2] | 56.36 [54.71-58.02] | 8.84 [8.3-9.37] | 59.14 [57.1-61.17] | 8.68 [8.19-9.16] |
| 2011 | 20.04 [19.03-21.05] | 12.65 [12.05-13.24] | 57.11 [55.35-58.88] | 9.08 [8.55-9.62] | 59.65 [57.19-62.1] | 8.85 [8.35-9.35] |
| 2012 | 19.59 [18.61-20.58] | 12.34 [11.74-12.94] | 56.01 [54.41-57.61] | 8.72 [8.23-9.21] | 58.72 [56.63-60.8] | 8.54 [8.06-9.02] |
| 2013 | 19.55 [18.52-20.58] | 12.31 [11.64-12.98] | 55.96 [53.97-57.95] | 8.7 [8.15-9.25] | 58.58 [56.08-61.09] | 8.5 [7.95-9.04] |
| 2014 | 18.28 [17.25-19.31] | 11.44 [10.86-12.02] | 54.1 [52.47-55.73] | 8.13 [7.65-8.61] | 56.78 [54.5-59.06] | 7.94 [7.49-8.39] |
| 2016 | 13.3 [12.62-13.98] | 8.14 [7.75-8.53] | 44.37 [42.88-45.87] | 5.65 [5.33-5.96] | 47.01 [44.93-49.1] | 5.51 [5.21-5.8] |
| 2017 | 13.59 [12.88-14.31] | 8.33 [7.91-8.75] | 45.6 [44.05-47.16] | 5.92 [5.6-6.24] | 47.48 [45.33-49.62] | 5.6 [5.3-5.91] |
| 2018 | 12.7 [11.9-13.49] | 7.75 [7.31-8.19] | 43.41 [41.6-45.21] | 5.44 [5.1-5.78] | 45.38 [42.89-47.86] | 5.17 [4.85-5.49] |
| 2019 | 12.23 [11.38-13.08] | 7.45 [6.95-7.94] | 42.57 [40.64-44.49] | 5.27 [4.88-5.66] | 43.83 [41.32-46.35] | 4.87 [4.51-5.24] |

[1] MDE: AAPC = -4.8 (95% CI, -6.1 to -3.6), $p < 0.001$

[2] No MDE: AAPC = -5.2 (95% CI, -6.5 to -3.8), $p < 0.001$

[3] AUD: AAPC = -2.7 (95% CI, -3.5 to -1.9), $p < 0.001$

[4] No AUD: AAPC = -5.1 (95% CI, -6.6 to -3.6), $p < 0.001$

[5] MDE and AUD: AAPC = -2.8 (95% CI, -3.4 to -2.1), $p < 0.001$

[6] No MDE or AUD: AAPC = -5.5 (95% CI, -6.8 to -4.1), $p < 0.001$

**B. Married**

| Year | MDE[1] | No MDE[2] | AUD[3] | No AUD[4] | MDE and AUD[5] | No MDE or AUD[6] |
|---|---|---|---|---|---|---|
| 2005 | 15.01 [14.25-15.77] | 9.26 [8.82-9.69] | 51.31 [49.77-52.86] | 7.33 [6.95-7.7] | 54.43 [52.33-56.54] | 7.27 [6.91-7.63] |
| 2006 | 14.36 [13.57-15.15] | 8.83 [8.4-9.25] | 49.98 [48.32-51.64] | 6.97 [6.6-7.35] | 53.07 [5.07-55.45] | 6.91 [6.55-7.27] |
| 2007 | 14.5 [13.71-15.28] | 8.92 [8.5-9.34] | 50.38 [48.62-52.13] | 7.08 [6.67-7.49] | 53.21 [50.79-55.62] | 6.95 [6.57-7.32] |
| 2008 | 14.3 [13.48-15.12] | 8.79 [8.32-9.26] | 49.86 [48.09-51.64] | 6.94 [6.56-7.33] | 52.79 [50.35-55.22] | 6.84 [6.46-7.22] |
| 2009 | 13.66 [12.93-14.39] | 8.37 [7.95-8.79] | 48.45 [46.71-50.18] | 6.59 [6.22-6.95] | 51.38 [49.13-53.63] | 6.49 [6.14-6.84] |
| 2010 | 12.97 [12.21-13.73] | 7.92 [7.52-8.24] | 47.32 [45.6-49.05] | 6.32 [5.92-6.71] | 50.03 [47.81-52.26] | 6.17 [5.82-6.52] |
| 2011 | 13.03 [12.27-13.8] | 7.97 [7.54-8.39] | 48.09 [46.24-49.93] | 6.5 [6.11-6.89] | 50.56 [47.87-53.25] | 6.29 [5.92-6.67] |
| 2012 | 12.72 [12.09-13.34] | 7.76 [7.41-8.11] | 46.97 [45.49-48.45] | 6.23 [5.92-6.54] | 49.6 [47.48-51.72] | 6.07 [5.77-6.37] |
| 2013 | 12.69 [11.89-13.49] | 7.74 [7.25-8.24] | 46.92 [44.86-48.97] | 6.22 [5.82-6.62] | 49.46 [46.75-52.17] | 6.04 [5.63-6.44] |
| 2014 | 11.8 [11.08-12.51] | 7.17 [6.79-7.55] | 45.05 [43.43-46.67] | 5.8 [5.46-6.13] | 47.62 [45.2-50.03] | 5.63 [5.31-5.95] |
| 2016 | 8.4 [7.96-8.84] | 5.03 [4.79-5.27] | 35.69 [34.33-37.05] | 4 [3.79-4.21] | 38.04 [36.01-40.07] | 3.87 [3.68-4.07] |
| 2017 | 8.6 [8.13-9.06] | 5.15 [4.89-5.41] | 36.84 [35.41-38.26] | 4.19 [3.98-4.4] | 38.48 [36.37-40.58] | 3.94 [3.74-4.15] |
| 2018 | 8 [7.46-8.53] | 4.78 [4.5-5.07] | 34.79 [33.1-36.49] | 3.85 [3.61-4.09] | 36.5 [34.09-38.91] | 3.64 [3.41-3.86] |
| 2019 | 7.69 [7.14-8.24] | 4.59 [4.28-4.9] | 34.02 [32.3-35.74] | 3.72 [3.46-3.99] | 35.06 [32.71-37.42] | 3.42 [3.18-3.67] |

[1] MDE: AAPC = -5.2 (95% CI, -6.5 to -3.8), $p < 0.001$

[2] No MDE: AAPC = -5.4 (95% CI, -6.8 to -4.0), $p < 0.001$

[3] AUD: AAPC= -3.2 (95% CI, -4.1 to -2.3), $p < 0.001$

[4] No AUD: AAPC = -5.2 (95% CI, -6.7 to -3.7), $p < 0.001$

[5] MDE and AUD: AAPC = -3.3 (95% CI, -4.1 to -2.5), $p < 0.001$

[6] No MDE or AUD: AAPC = -5.6 (95% CI, -7.0 to -4.2), $p < 0.001$

**C. Widowed**

| Year | MDE[1] | No MDE[2] | AUD[3] | No AUD[4] | MDE and AUD[5] | No MDE or AUD[6] |
|---|---|---|---|---|---|---|
| 2005 | 21.38 [20.18-22.58] | 13.57 [12.82-14.33] | 59.66 [57.9-61.43] | 9.99 [9.39-10.59] | 61.73 [59.62-63.84] | 9.58 [9.01-10.14] |
| 2006 | 20.51 [19.39-21.64] | 12.97 [12.32-13.62] | 58.37 [56.69-60.06] | 9.52 [9-10.04] | 60.43 [58.18-62.68] | 9.11 [8.62-9.61] |
| 2007 | 20.7 [19.48-21.91] | 13.1 [12.38-13.82] | 58.76 [56.82-60.7] | 9.66 [9.03-10.29] | 60.56 [58.17-62.94] | 9.16 [8.59-9.72] |
| 2008 | 20.44 [19.25-21.62] | 12.92 [12.19-13.64] | 58.26 [56.28-60.24] | 9.48 [8.87-10.09] | 60.15 [57.75-62.55] | 9.02 [8.46-9.58] |
| 2009 | 19.59 [18.55-20.62] | 12.33 [11.71-12.95] | 56.88 [55.12-58.64] | 9.01 [8.51-9.5] | 58.8 [56.66-60.93] | 8.57 [8.09-9.04] |
| 2010 | 18.66 [17.55-19.77] | 11.7 [11.07-12.33] | 55.77 [53.9-57.64] | 8.64 [8.07-9.22] | 57.48 [55.26-59.71] | 8.15 [7.64-8.67] |
| 2011 | 18.74 [17.64-19.85] | 11.76 [11.1-12.41] | 56.52 [54.56-58.49] | 8.89 [8.32-9.46] | 58 [55.36-60.63] | 8.31 [7.78-8.85] |
| 2012 | 18.32 [17.24-19.39] | 11.47 [10.81-12.12] | 55.42 [53.58-57.25] | 8.53 [8-9.07] | 57.06 [54.81-59.31] | 8.02 [7.52-8.53] |
| 2013 | 18.28 [17.18-19.38] | 11.44 [10.74-12.14] | 55.36 [53.23-57.5] | 8.51 [7.95-9.08] | 56.92 [54.29-59.56] | 7.98 [7.44-8.53] |
| 2014 | 17.07 [16.06-18.09] | 10.63 [10.07-11.18] | 53.5 [51.76-55.25] | 7.95 [7.47-8.43] | 55.1 [52.75-57.46] | 7.46 [7.02-7.89] |
| 2016 | 12.37 [11.64-13.1] | 7.54 [7.12-7.95] | 43.78 [42.04-45.52] | 5.55 [5.18-5.87] | 45.32 [43.09-47.56] | 5.16 [4.85-5.47] |
| 2017 | 12.65 [11.88-13.41] | 7.72 [7.27-8.16] | 45.01 [43.24-46.78] | 5.79 [5.44-6.13] | 45.78 [43.5-48.07] | 5.25 [4.93-5.57] |
| 2018 | 11.8 [10.99-12.61] | 7.17 [6.72-7.62] | 42.82 [40.8-44.84] | 5.32 [4.95-5.69] | 43.7 [41.09-46.3] | 4.85 [4.52-5.18] |
| 2019 | 11.36 [10.53-12.2] | 6.89 [6.41-7.38] | 41.98 [39.95-44.02] | 5.15 [4.76-5.54] | 42.17 [39.58-44.75] | 4.57 [4.21-4.93] |

[1] MDE: AAPC = -4.8 (95% CI, -6.1 to -3.6), $p < 0.001$

[2] No MDE: AAPC = -5.2 (95% CI, -6.5 to -3.9), $p < 0.001$

[3] AUD: AAPC = -2.7 (95% CI, -3.4 to -2.0), $p < 0.001$

[4] No AUD: AAPC = -5.1 (95% CI, -6.5 to -3.6), $p < 0.001$

[5] MDE and AUD: AAPC = -2.8 (95% CI, -3.5 to -2.2), $p < 0.001$

[6] No MDE or AUD: AAPC = -5.5 (95% CI, -6.7 to -4.2), $p < 0.001$

**Supplemental Table 6.** Trends in past-year adjusted prevalence of driving under the influence of alcohol (DUIA) and 95% CI by past-year major depressive episode (MDE) and alcohol use disorder (AUD), stratified by education level, from 2005 to 2019.

**A. Less than high school**

| Year | MDE[1] | No MDE[2] | AUD[3] | No AUD[4] | MDE and AUD[5] | No MDE or AUD[6] |
|---|---|---|---|---|---|---|
| 2005 | 10.73 [10.02-11.44] | 6.49 [6.08-6.9] | 38.04 [36.23-39.86] | 4.4 [4.09-4.72] | 42.34 [40.1-44.58] | 4.6 [4.29-4.91] |
| 2006 | 10.24 [9.54-10.94] | 6.18 [5.8-6.56] | 36.8 [35.07-38.52] | 4.19 [3.9-4.47] | 41.01 [38.62-43.4] | 4.37 [4.08-4.66] |
| 2007 | 10.35 [9.62-11.97] | 6.25 [5.85-6.65] | 37.17 [35.19-39.15] | 4.25 [3.92-4.58] | 41.14 [38.61-43.68] | 4.39 [4.07-4.71] |
| 2008 | 10.2 [9.55-10.85] | 6.15 [5.79-6.52] | 36.69 [34.98-38.4] | 4.17 [3.9-4.44] | 40.73 [38.42-43.05] | 4.32 [4.05-4.58] |
| 2009 | 9.72 [9.04-10.4] | 5.86 [5.46-6.25] | 35.38 [33.48-37.28] | 3.95 [3.65-4.24] | 39.38 [37.03-41.73] | 4.09 [3.79-4.39] |
| 2010 | 9.21 [8.55-9.86] | 5.53 [5.18-5.89] | 34.36 [32.56-36.16] | 3.78 [3.49-4.07] | 38.1 [35.88-40.33] | 3.88 [3.6-4.16] |
| 2011 | 9.25 [8.59-9.92] | 5.56 [5.18-5.94] | 35.05 [33.13-36.97] | 3.89 [3.59-4.19] | 38.6 [35.94-41.26] | 3.96 [3.67-4.26] |
| 2012 | 9.02 [8.4-9.64] | 5.42 [5.06-5.77] | 34.04 [32.37-35.71] | 3.73 [3.46-3.99] | 37.69 [35.48-39.91] | 3.82 [3.55-4.09] |
| 2013 | 9 [8.35-9.65] | 5.4 [5.02-5.79] | 33.99 [32.06-35.92] | 3.72 [3.44-3.99] | 37.56 [35.03-40.1] | 3.8 [3.52-4.08] |
| 2014 | 8.34 [7.73-8.95] | 4.99 [4.67-5.32] | 32.33 [30.7-33.95] | 3.46 [3.21-3.71] | 35.85 [33.53-38.17] | 3.54 [3.29-3.79] |
| 2016 | 5.87 [5.47-6.28] | 3.48 [3.26-3.7] | 24.43 [23.03-25.83] | 2.37 [2.19-2.54] | 27.4 [25.53-29.27] | 2.42 [2.25-2.59] |
| 2017 | 6.02 [5.6-6.43] | 3.57 [3.33-3.8] | 25.36 [23.96-26.76] | 2.49 [2.31-2.66] | 27.77 [25.87-29.67] | 2.46 [2.29-2.63] |
| 2018 | 5.59 [5.14-6.03] | 3.3 [3.06-3.55] | 23.71 [22.17-25.25] | 2.28 [2.1-2.46] | 26.11 [23.98-28.24] | 2.27 [2.09-2.45] |
| 2019 | 5.36 [4.89-5.84] | 3.17 [2.91-3.43] | 23.1 [21.45-24.75] | 2.2 [2-2.41] | 24.92 [22.8-27.05] | 2.13 [1.94-2.33] |

[1] MDE: AAPC = -5.3 (95% CI, -6.7 to -3.9), $p < 0.001$

[2] No MDE: AAPC = -5.5 (95% CI, -6.9 to -4.1), $p < 0.001$

[3] AUD: AAPC = -3.8 (95% CI, -4.9 to -2.8), $p < 0.001$

[4] No AUD: AAPC = -4.5 (95% CI, -6.4 to -2.6), $p < 0.001$

[5] MDE and AUD: AAPC = -4.0 (95% CI, -4.9 to -3.0), $p < 0.001$

[6] No MDE or AUD: AAPC = -5.8 (95% CI, -7.2 to -4.3), $p < 0.001$

**B. High school diploma**

| Year | MDE[1] | No MDE[2] | AUD[3] | No AUD[4] | MDE and AUD[5] | No MDE or AUD[6] |
|---|---|---|---|---|---|---|
| 2005 | 15.72 [14.91-16.53] | 9.73 [9.27-10.18] | 51.12 [49.52-52.72] | 7.28 [6.88-7.67] | 54.27 [52.2-56.33] | 7.23 [6.86-7.59] |
| 2006 | 15.04 [14.19-15.88] | 9.27 [8.82-9.73] | 49.79 [48.14-51.44] | 6.92 [6.55-7.3] | 52.91 [50.6-55.22] | 6.87 [6.51-7.23] |
| 2007 | 15.18 [14.33-16.04] | 9.37 [8.91-9.83] | 50.19 [48.42-51.96] | 7.03 [6.61-7.44] | 53.04 [50.66-55.42] | 6.9 [6.52-7.29] |
| 2008 | 14.98 [14.11-15.84] | 9.24 [8.74-9.73] | 49.67 [47.83-51.51] | 6.9 [6.48-7.31] | 52.62 [50.2-55.04] | 6.8 [6.41-7.19] |
| 2009 | 14.31 [13.55-15.07] | 8.8 [8.37-9.23] | 48.26 [46.54-49.97] | 6.54 [6.18-6.9] | 51.21 [49.02-53.41] | 6.45 [6.09-6.8] |
| 2010 | 13.6 [12.78-14.41] | 8.33 [7.9-8.76] | 47.13 [45.43-48.84] | 6.27 [5.88-6.66] | 49.86 [47.71-52.02] | 6.13 [5.78-6.48] |
| 2011 | 13.66 [12.87-14.46] | 8.37 [7.93-8.81] | 47.9 [46.08-49.72] | 6.45 [6.06-6.84] | 50.39 [47.76-53.02] | 6.25 [5.88-6.63] |
| 2012 | 13.33 [12.66-14.01] | 8.16 [7.79-8.53] | 46.78 [45.23-48.32] | 6.19 [5.86-6.52] | 49.43 [47.35-51.52] | 6.03 [5.72-6.34] |
| 2013 | 13.3 [12.48-14.12] | 8.14 [7.64-8.64] | 46.72 [44.66-48.79] | 6.17 [5.8-6.58] | 49.29 [46.62-51.96] | 6 [5.59-6.41] |
| 2014 | 12.37 [11.6-13.15] | 7.54 [7.13-7.95] | 44.86 [43.17-46.56] | 5.75 [5.4-6.11] | 47.45 [45.05-49.85] | 5.6 [5.27-5.93] |
| 2016 | 8.83 [8.35-9.31] | 5.3 [5.04-5.56] | 35.51 [34.13-36.89] | 3.97 [3.75-4.18] | 37.88 [35.88-39.88] | 3.85 [3.65-4.05] |
| 2017 | 9.03 [8.54-9.53] | 5.42 [5.15-5.7] | 36.66 [35.23-38.08] | 4.16 [3.95-4.38] | 38.32 [36.27-40.37] | 3.92 [3.71-4.12] |
| 2018 | 8.41 [7.83-8.98] | 5.03 [4.73-5.34] | 34.62 [32.87-36.37] | 3.82 [3.57-4.07] | 36.34 [33.95-38.74] | 3.63 [3.38-3.85] |
| 2019 | 8.08 [7.49-8.67] | 4.83 [4.51-5.16] | 33.85 [32.08-35.61] | 3.7 [3.42-3.97] | 34.91 [32.56-37.26] | 3.4 [3.15-3.66] |

[1] MDE: AAPC = -5.1 (95% CI, -6.5 to -3.8), $p < 0.001$

[2] No MDE: AAPC = -5.4 (95% CI, -6.7 to -4.0), $p < 0.001$

[3] AUD: AAPC = -3.2 (95% CI, -4.1 to -2.3), $p < 0.001$

[4] No AUD: AAPC = -5.2 (95% CI, -6.8 to -3.7), $p < 0.001$

[5] MDE and AUD: AAPC = -3.3 (95% CI, -4.1 to -2.5), $p < 0.001$

[6] No MDE or AUD: AAPC = -5.7 (95% CI, -7.1 to -4.2), $p < 0.001$

**C. Some college**

| Year | MDE[1] | No MDE[2] | AUD[3] | No AUD[4] | MDE and AUD[5] | No MDE or AUD[6] |
|---|---|---|---|---|---|---|
| 2005 | 20.18 [19.2-21.17] | 12.74 [12.13-13.36] | 58.93 [57.33-60.53] | 9.72 [9.19-10.25] | 61.18 [59.16-63.2] | 9.38 [8.88-9.88] |
| 2006 | 19.35 [18.39-20.31] | 12.17 [11.63-12.72] | 57.64 [56.03-59.24] | 9.26 [8.78-9.75] | 59.88 [57.67-62.08] | 8.92 [8.47-9.38] |
| 2007 | 19.53 [18.55-20.51] | 12.3 [11.73-12.86] | 58.03 [56.29-59.76] | 9.4 [8.86-9.93] | 60.01 [57.75-62.26] | 8.97 [8.49-9.44] |
| 2008 | 19.28 [18.27-20.29] | 12.12 [11.5-12.74] | 57.52 [55.78-59.27] | 9.22 [8.72-9.73] | 59.6 [57.3-61.89] | 8.33 [8.35-9.31] |
| 2009 | 18.47 [17.6-19.34] | 11.57 [11.04-12.09] | 56.13 [54.53-57.73] | 8.76 [8.33-9.19] | 58.24 [56.23-60.24] | 8.39 [7.99-8.79] |
| 2010 | 17.58 [16.64-18.52] | 10.97 [10.45-11.49] | 55.02 [53.38-56.67] | 8.41 [7.92-8.9] | 56.92 [54.85-58.98] | 7.98 [7.55-8.41] |
| 2011 | 17.66 [16.73-18.59] | 11.02 [10.48-11.57] | 55.78 [53.94-57.62] | 8.65 [8.13-9.16] | 57.43 [54.9-59.96] | 8.14 [7.68-8.6] |
| 2012 | 17.26 [16.44-18.07] | 10.75 [10.26-11.24] | 54.67 [53.08-56.25] | 8.3 [7.85-8.74] | 56.49 [54.42-58.57] | 7.86 [7.44-8.27] |
| 2013 | 17.22 [16.26-18.18] | 10.73 [10.1-11.35] | 54.61 [52.6-56.63] | 8.28 [7.77-8.8] | 56.36 [53.79-58.93] | 7.82 [7.31-8.32] |
| 2014 | 16.07 [15.18-16.95] | 9.96 [9.47-10.44] | 52.75 [51.15-54.35] | 7.73 [7.3-8.16] | 54.53 [52.23-56.83] | 7.3 [6.91-7.69] |
| 2016 | 11.6 [11.02-12.19] | 7.05 [6.71-7.39] | 43.04 [41.54-44.53] | 5.36 [5.08-5.65] | 44.75 [42.66-46.84] | 5.05 [4.79-5.31] |
| 2017 | 11.87 [11.27-12.46] | 7.22 [6.87-7.56] | 44.26 [42.74-45.78] | 5.63 [5.34-5.9] | 45.21 [43.09-47.33] | 5.14 [4.88-5.4] |
| 2018 | 11.07 [10.38-11.76] | 6.71 [6.32-7.09] | 42.08 [40.26-43.9] | 5.17 [4.85-5.49] | 43.13 [40.66-45.6] | 4.74 [4.46-5.03] |
| 2019 | 10.65 [9.93-11.37] | 6.44 [6.03-6.86] | 41.25 [39.41-43.09] | 5 [4.66-5.35] | 41.6 [39.17-44.04] | 4.47 [4.15-4.79] |

[1] MDE: AAPC = -4.9 (95% CI, -6.2 to -3.6), $p < 0.005$

[2] No MDE: AAPC = -5.2 (95% CI, -6.6 to -3.9), $p < 0.001$

[3] AUD: AAPC = -2.8 (95% CI, -3.5 to -2.0), $p < 0.001$

[4] No AUD: AAPC = -5.1 (95% CI, -6.6 to -3.6), $p < 0.001$

[5] MDE and AUD: AAPC = -2.9 (95% CI, -3.5 to -2.2), $p < 0.001$

[6] No MDE or AUD: AAPC = -5.5 (95% CI, -6.9 to -4.1), $p < 0.001$

**D. College graduate**

| Year | MDE[1] | No MDE[2] | AUD[3] | No AUD[4] | MDE and AUD[5] | No MDE or AUD[6] |
|---|---|---|---|---|---|---|
| 2005 | 23.42 [22.31-24.52] | 15.01 [14.33-15.69] | 64.15 [62.68-65.62] | 11.84 [11.25-12.42] | 66.58 [64.66-68.5] | 11.57 [11.02-12.12] |
| 2006 | 22.49 [21.39-23.59] | 14.35 [13.73-14.98] | 62.92 [61.36-64.47] | 11.29 [10.72-11.87] | 65.35 [63.21-67.5] | 11.02 [10.49-11.55] |
| 2007 | 22.69 [21.55-23.83] | 14.49 [13.84-15.15] | 63.29 [61.63-64.94] | 11.45 [10.82-12.08] | 65.48 [63.27-67.68] | 11.07 [10.51-11.64] |
| 2008 | 22.41 [21.2-23.62] | 14.3 [13.54-15.05] | 62.81 [61.01-64.61] | 11.25 [10.59-11.91] | 65.09 [62.79-67.39] | 10.91 [10.29-11.52] |
| 2009 | 21.5 [20.47-22.52] | 13.66 [13.04-14.27] | 61.47 [59.8-63.15] | 10.69 [10.12-11.26] | 63.8 [61.72-65.89] | 10.37 [9.84-10.9] |
| 2010 | 20.51 [19.4-21.61] | 12.97 [12.35-13.58] | 60.4 [58.68-62.13] | 10.27 [9.64-10.9] | 62.55 [60.43-64.67] | 9.88 [9.33-10.43] |
| 2011 | 20.6 [19.54-21.66] | 13.03 [12.41-13.65] | 61.13 [59.39-62.88] | 10.56 [9.96-11.15] | 63.04 [60.55-65.54] | 10.07 [9.51-10.63] |
| 2012 | 20.14 [19.15-21.13] | 12.71 [12.12-13.3] | 60.06 [58.54-61.57] | 10.14 [9.63-10.65] | 62.14 [60.06-64.22] | 9.73 [9.23-10.22] |
| 2013 | 20.1 [18.97-21.23] | 12.68 [11.95-13.42] | 60.01 [57.99-62.03] | 10.12 [9.48-10.76] | 62.01 [59.44-64.58] | 9.68 [9.06-10.31] |
| 2014 | 18.8 [17.78-19.81] | 11.79 [11.24-12.34] | 58.2 [56.62-59.77] | 9.46 [8.95-9.97] | 60.26 [57.98-62.54] | 9.05 [8.59-9.51] |
| 2016 | 13.7 [13.03-14.37] | 8.4 [8.02-8.77] | 48.51 [47.02-50] | 6.6 [6.27-6.94] | 50.59 [48.44-52.74] | 6.3 [6-6.6] |
| 2017 | 14 [13.25-14.75] | 8.59 [8.16-9.03] | 49.75 [48.12-51.38] | 6.92 [6.56-7.28] | 51.05 [48.77-53.34] | 6.41 [6.07-6.75] |
| 2018 | 13.08 [12.28-13.88] | 8 [7.56-8.43] | 47.53 [45.7-49.36] | 6.36 [5.99-6.74] | 48.95 [46.37-51.52] | 5.92 [5.58-6.26] |
| 2019 | 12.6 [11.77-13.43] | 7.69 [7.21-8.16] | 46.68 [44.8-48.56] | 6.16 [5.75-6.58] | 47.38 [44.83-49.94] | 5.58 [5.2-5.96] |

[1] MDE: AAPC = -4.8 (95% CI, -6.0 to -3.5), $p < 0.001$

[2] No MDE: AAPC= -5.1 (95% CI, -6.5 to -3.8), $p < 0.001$

[3] AUD: AAPC = -2.5 (95% CI, -3.2 to -1.8), $p < 0.001$

[4] No AUD: AAPC = -5.0 (95% CI, -6.5 to -3.5), $p < 0.001$

[5] MDE and AUD: AAPC = -2.5 (95% CI, -3.1 to -2.0), $p < 0.001$

[6] No MDE or AUD: AAPC= -5.5 (95% CI, -6.8 to -4.1), $p < 0.001$

**Supplemental Table 7.** Trends in past-year adjusted prevalence of driving under the influence of alcohol (DUIA) and 95% CI by past-year major depressive episode (MDE) and alcohol use disorder (AUD), stratified by income, from 2005 to 2019.

**A. Less than $20,000**

| Year | MDE[1] | No MDE[2] | AUD[3] | No AUD[4] | MDE and AUD[5] | No MDE or AUD[6] |
|---|---|---|---|---|---|---|
| 2005 | 13.22 [12.5-13.93] | 8.08 [7.67-8.5] | 43.93 [42.28-45.58] | 5.55 [5.24-5.87] | 46.41 [44.28-48.54] | 5.38 [5.08-5.68] |
| 2006 | 12.63 [11.88-13.37] | 7.7 [7.29-8.12] | 42.62 [40.8-44.45] | 5.28 [4.94-5.61] | 45.05 [42.65-48.45] | 5.11 [4.81-5.41] |
| 2007 | 12.75 [12.02-13.48] | 7.78 [7.38-8.19] | 43.01 [41.25-44.78] | 5.36 [5.04-5.68] | 45.19 [42.8-47.57] | 5.13 [4.84-5.43] |
| 2008 | 12.57 [11.81-13.34] | 7.67 [7.22-8.12] | 42.51 [40.59-44.43] | 5.26 [4.92-5.6] | 44.77 [42.31-47.23] | 5.05 [4.74-5.37] |
| 2009 | 12 [11.35-12.65] | 7.3 [6.93-7.68] | 41.13 [39.39-42.87] | 4.98 [4.69-5.27] | 43.38 [41.22-45.54] | 4.79 [4.52-5.06] |
| 2010 | 11.38 [10.71-12.06] | 6.91 [6.55-7.26] | 40.05 [38.33-41.76] | 4.77 [4.46-5.08] | 42.06 [39.93-44.19] | 4.55 [4.28-4.82] |
| 2011 | 11.44 [10.76-12.12] | 6.94 [6.56-7.33] | 40.78 [38.91-42.66] | 4.91 [4.59-5.23] | 42.57 [39.97-45.18] | 4.64 [4.35-4.93] |
| 2012 | 11.16 [10.57-11.74] | 6.76 [6.43-7.1] | 39.7 [38.15-41.25] | 4.71 [4.44-4.97] | 41.64 [39.55-43.72] | 4.47 [4.23-4.72] |
| 2013 | 11.13 [10.39-11.87] | 6.75 [6.29-7.2] | 39.65 [37.54-41.77] | 4.7 [4.36-5.04] | 41.5 [38.84-44.16] | 4.45 [4.13-4.77] |
| 2014 | 10.34 [9.67-11.01] | 6.24 [5.88-6.6] | 37.87 [36.14-39.6] | 4.37 [4.08-4.67] | 39.72 [37.36-42.08] | 4.15 [3.89-4.41] |
| 2016 | 7.33 [6.92-8.77] | 4.37 [4.14-4.59] | 29.21 [27.88-30.53] | 3 [2.83-3.18] | 30.8 [28.95-32.65] | 2.84 [2.68-3] |
| 2017 | 7.5 [7.06-7.94] | 4.47 [4.22-4.73] | 30.24 [28.82-31.67] | 3.15 [2.97-3.33] | 31.19 [29.93-33.14] | 2.89 [2.72-3.06] |
| 2018 | 6.97 [6.5-7.44] | 4.15 [3.9-4.4] | 28.4 [26.84-29.96] | 2.89 [2.7-3.08] | 29.41 [27.29-31.54] | 2.66 [2.49-2.83] |
| 2019 | 6.7 [6.21-7.19] | 3.98 [3.71-4.26] | 27.71 [26.09-29.33] | 2.8 [2.58-3.01] | 28.13 [26.03-30.24] | 2.51 [2.31-2.7] |

[1] MDE: AAPC = -5.2 (95% CI, -6.5 to -3.9), $p < 0.001$

[2] No MDE: AAPC = -5.5 (95% CI, -6.9 to -4.0), $p < 0.001$

[3] AUD: AAPC = -3.6 (95% CI, -4.6 to -2.5), $p < 0.001$

[4] No AUD: AAPC = -5.3 (95% CI, -6.8 to -3.7), $p < 0.001$

[5] MDE and AUD: AAPC= -3.7 (95% CI, -4.6 to -2.9), $p < 0.001$

[6] No MDE or AUD: AAPC = -5.7 (95% CI, -7.1 to -4.3), $p < 0.001$

**B. $20,000-$49,999**

| Year | MDE[1] | No MDE[2] | AUD[3] | No AUD[4] | MDE and AUD[5] | No MDE or AUD[6] |
|---|---|---|---|---|---|---|
| 2005 | 16.14 [15.31-16.97] | 10 [9.52-10.48] | 51.96 [50.36-53.57] | 7.51 [7.09-7.92] | 54.67 [52.57-56.77] | 7.34 [6.96-7.72] |
| 2006 | 15.44 [14.61-16.27] | 9.54 [9.1-9.98] | 50.63 [49.03-52.23] | 7.15 [6.76-7.53] | 53.31 [51-55.62] | 6.97 [6.62-7.33] |
| 2007 | 15.59 [14.78-16.4] | 9.64 [9.21-10.07] | 51.03 [49.36-52.69] | 7.25 [6.85-7.65] | 53.45 [51.13-55.76] | 7 [6.66-7.36] |
| 2008 | 15.38 [14.5-16.26] | 9.5 [8.99-10.1] | 50.52 [48.73-52.3] | 7.11 [6.7-7.53] | 53.02 [50.61-55.43] | 6.9 [6.51-7.29] |
| 2009 | 14.7 [13.91-15.5] | 9.05 [8.6-9.51] | 49.1 [47.37-50.83] | 6.75 [6.37-7.13] | 51.62 [49.4-53.84] | 6.55 [6.19-6.91] |
| 2010 | 13.97 [13.15-14.78] | 8.57 [8.14-9] | 47.97 [46.25-49.7] | 6.47 [6.06-6.88] | 50.27 [48.07-52.47] | 6.22 [5.86-6.59] |
| 2011 | 14.03 [13.22-14.84] | 8.62 [8.16-9.07] | 48.74 [46.89-50.59] | 6.66 [6.25-7.07] | 50.8 [48.13-53.47] | 6.35 [5.96-6.74] |
| 2012 | 13.7 [13.02-14.37] | 8.4 [8.02-8.77] | 47.62 [46.15-49.09] | 6.39 [6.06-6.71] | 49.84 [47.75-51.93] | 6.12 [5.82-6.43] |
| 2013 | 13.67 [12.8-14.53] | 8.38 [7.84-8.91] | 47.56 [45.48-49.65] | 6.37 [5.94-6.8] | 49.7 [46.99-52.41] | 6.09 [5.67-6.51] |
| 2014 | 12.72 [11.95-13.48] | 7.76 [7.36-8.16] | 45.7 [44.09-47.3] | 5.94 [5.59-6.29] | 47.86 [45.48-50.23] | 5.68 [5.36-6] |
| 2016 | 9.08 [8.6-9.57] | 5.45 [5.19-5.72] | 36.29 [34.91-37.66] | 4.1 [3.87-4.32] | 38.26 [36.24-40.29] | 3.91 [3.71-4.11] |
| 2017 | 9.29 [8.78-9.08] | 5.59 [5.3-5.87] | 37.44 [36.02-38.86] | 4.3 [4.08-4.52] | 38.7 [36.62-40.79] | 3.98 [3.77-4.19] |
| 2018 | 8.65 [8.07-9.23] | 5.19 [4.87-5.5] | 35.38 [33.69-37.08] | 3.95 [3.7-4.2] | 36.72 [34.33-39.11] | 3.67 [3.44-3.9] |
| 2019 | 8.32 [7.71-8.92] | 4.98 [4.64-4.32] | 34.61 [32.87-36.35] | 3.82 [3.54-4.1] | 35.28 [32.93-37.63] | 3.46 [3.2-3.71] |

[1] MDE: AAPC = -5.1 (95% CI, -6.4 to -3.8), $p < 0.001$

[2] No MDE: AAPC = -5.4 (95% CI, -6.8 to -4.0), $p < 0.001$

[3] AUD: AAPC = -3.2 (95% CI, -4.1 to -2.3), $p < 0.001$

[4] No AUD: AAPC = -5.2 (95% CI, -6.8 to -3.7), $p < 0.001$

[5] MDE and AUD: AAPC = -3.3 (95% CI, -4.0 to -2.5), $p < 0.001$

[6] No MDE or AUD: AAPC = -5.7 (95% CI, -7.1 to -4.2), $p < 0.001$

**C. $50,00-$74,999**

| Year | MDE[1] | No MDE[2] | AUD[3] | No AUD[4] | MDE and AUD[5] | No MDE or AUD[6] |
|---|---|---|---|---|---|---|
| 2005 | 19.65 [18.54-20.76] | 12.37 [11.68-13.07] | 58.92 [57.12-60.72] | 9.72 [9.12-10.32] | 61.56 [59.48-63.64] | 9.52 [8.95-10.08] |
| 2006 | 18.83 [17.82-19.85] | 11.82 [11.23-12.4] | 57.63 [55.95-59.3] | 9.26 [8.75-9.76] | 60.26 [58.07-62.45] | 9.06 [8.58-9.53] |
| 2007 | 19.01 [17.9-20.11] | 11.94 [11.28-12.59] | 58.01 [56.04-59.99] | 9.39 [8.77-10.18] | 60.39 [58.03-62.75] | 9.1 [8.54-9.66] |
| 2008 | 18.76 [17.73-19.79] | 11.77 [11.14-12.4] | 57.51 [55.7-59.33] | 9.22 [8.69-9.75] | 59.98 [57.7-62.27] | 8.96 [8.45-9.47] |
| 2009 | 17.97 [17.01-18.92] | 11.23 [10.65-11.81] | 56.12 [54.32-57.93] | 8.76 [8.26-9.26] | 58.62 [56.5-60.75] | 8.51 [8.03-9] |
| 2010 | 17.1 [16.05-18.15] | 10.64 [10.04-11.25] | 55.01 [53.12-56.9] | 8.4 [7.84-8.97] | 57.31 [55.13-59.49] | 8.1 [7.59-8.61] |
| 2011 | 17.18 [16.22-18.14] | 10.7 [10.13-11.27] | 55.77 [53.9-57.63] | 8.64 [8.12-9.16] | 57.82 [55.3-60.35] | 8.26 [7.77-8.75] |
| 2012 | 16.78 [15.83-17.73] | 10.43 [9.85-11.01] | 54.66 [52.86-56.45] | 8.29 [7.79-8.8] | 56.88 [54.7-59.06] | 7.97 [7.49-8.46] |
| 2013 | 16.74 [15.81-17.68] | 10.41 [9.81-11] | 54.5 [52.56-56.65] | 8.28 [7.76-8.8] | 56.75 [54.23-59.27] | 7.93 [7.42-8.44] |
| 2014 | 15.62 [14.7-16.53] | 9.66 [9.15-10.16] | 52.74 [51.08-54.4] | 7.73 [7.28-8.17] | 54.93 [52.66-57.2] | 7.41 [7-7.82] |
| 2016 | 11.26 [10.62-11.91] | 6.83 [6.46-6.2] | 43.03 [41.35-44.71] | 5.36 [5.04-5.69] | 45.15 [43-47.29] | 5.13 [4.83-5.42] |
| 2017 | 11.52 [10.87-12.16] | 6.99 [6.62-7.37] | 44.25 [42.58-45.92] | 5.62 [5.31-5.94] | 45.61 [43.43-47.78] | 5.22 [4.92-5.52] |
| 2018 | 10.74 [9.99-11.49] | 6.5 [6.08-6.92] | 42.07 [40.06-44.08] | 5.17 [4.81-5.53] | 43.52 [40.96-46.08] | 4.82 [4.49-5.14] |
| 2019 | 10.34 [9.56-11.12] | 6.24 [5.79-6.7] | 41.24 [39.15-43.32] | 5 [4.61-5.4] | 41.99 [39.43-44.55] | 4.54 [4.18-4.9] |

[1] MDE: AAPC = -4.9 (95% CI, -6.2 to -3.7), $p < 0.001$

[2] No MDE: AAPC = -5.2 (95% CI, -6.6 to -3.9), $p < 0.001$

[3] AUD: AAPC = -2.8 (95% CI, -3.5 to -2.0), $p < 0.001$

[4] No AUD: AAPC = -5.1 (95% CI, -6.6 to -3.6), $p < 0.001$

[5] MDE and AUD: AAPC = -2.9 (95% CI, -3.5 to -2.2), $p < 0.001$

[6] No MDE or AUD: AAPC = -5.5 (95% CI, -6.9 to -4.2), $p < 0.001$

**D. ≥ $75,000**

| Year | MDE[1] | No MDE[2] | AUD[3] | No AUD[4] | MDE and AUD[5] | No MDE or AUD[6] |
|---|---|---|---|---|---|---|
| 2005 | 22.51 [21.45-23.57] | 14.37 [13.74-15] | 62.63 [61.18-64.08] | 11.17 [10.62-11.72] | 65.91 [64.02-67.8] | 11.27 [10.73-11.8] |
| 2006 | 21.61 [20.51-22.7] | 13.73 [13.12-14.35] | 61.37 [59.86-62.88] | 10.65 [10.12-11.18] | 64.67 [62.55-66.8] | 10.73 [10.22-11.25] |
| 2007 | 21.8 [20.65-22.96] | 13.87 [13.2-14.54] | 61.75 [60.01-63.49] | 10.8 [10.16-11.44] | 64.8 [62.55-67.05] | 10.78 [10.19-11.38] |
| 2008 | 21.53 [20.41-22.65] | 13.68 [13-14.36] | 61.26 [59.55-62.98] | 10.61 [10.02-11.19] | 64.41 [62.18-66.64] | 10.62 [10.06-11.18] |
| 2009 | 20.65 [19.64-21.65] | 13.06 [12.47-13.66] | 59.91 [58.27-61.54] | 10.08 [9.56-10.6] | 63.11 [61.06-65.16] | 10.1 [9.59-10.61] |
| 2010 | 19.68 [18.61-20.75] | 12.4 [11.81-12.98] | 58.82 [57.2-60.44] | 9.68 [9.12-10.24] | 61.84 [59.8-63.89] | 9.62 [9.11-10.13] |
| 2011 | 19.77 [18.71-20.83] | 12.46 [11.84-13.08] | 59.56 [57.8-61.32] | 9.95 [9.38-10.52] | 62.34 [59.85-64.83] | 9.8 [9.25-10.35] |
| 2012 | 19.33 [18.35-20.3] | 12.15 [11.58-12.73] | 58.47 [56.92-60.02] | 9.56 [9.05-10.06] | 61.43 [59.37-63.5] | 9.47 [8.98-9.96] |
| 2013 | 19.29 [18.23-20.34] | 12.13 [11.45-12.8] | 58.42 [56.5-60.35] | 9.54 [8.97-10.1] | 61.3 [58.81-63.8] | 9.42 [8.85-10] |
| 2014 | 18.03 [17.02-19.03] | 11.27 [10.73-11.81] | 56.59 [55.02-58.16] | 8.91 [8.42-9.4] | 59.54 [57.25-61.82] | 8.81 [8.35-9.27] |
| 2016 | 13.1 [12.44-13.77] | 8.01 [7.64-8.38] | 46.87 [45.39-48.35] | 6.21 [5.89-6.53] | 49.84 [47.71-51.98] | 6.13 [5.83-6.42] |
| 2017 | 13.4 [12.69-14.1] | 8.2 [7.8-8.86] | 48.11 [46.56-49.66] | 6.5 [6.18-6.83] | 50.31 [48.1-52.52] | 6.23 [5.92-6.55] |
| 2018 | 12.51 [11.72-13.29] | 7.63 [7.2-8.05] | 45.9 [44.06-47.73] | 5.98 [5.62-6.35] | 48.2 [45.63-50.77] | 5.76 [5.41-6.1] |
| 2019 | 12.05 [11.24-12.85] | 7.33 [6.87-7.79] | 45.05 [43.19-46.91] | 5.79 [5.4-6.19] | 46.64 [44.1-49.18] | 5.43 [5.05-5.8] |

[1] MDE: AAPC = -4.8 (95% CI, -6.0 to -3.6), $p < 0.001$

[2] No MDE: AAPC = -5.2 (95% CI, -6.5 to -3.8), $p < 0.001$

[3] AUD: AAPC = -2.6 (95% CI, -3.3 to -1.8), $p < 0.001$

[4] No AUD: AAPC = -5.1 (95% CI, -6.6 to -3.6), $p < 0.001$

[5] MDE and AUD: AAPC = -2.6 (95% CI, -3.2 to -2.0), $p < 0.001$

[6] No MDE or AUD: AAPC = -5.5 (95% CI, -6.8 to -4.1), $p < 0.001$

**Supplemental Table 8.** Trends in past-year adjusted prevalence of driving under the influence of alcohol (DUIA) and 95% CI by past-year major depressive episode (MDE) and alcohol use disorder (AUD), stratified by metropolitan status, from 2005 to 2019.

**A. Large metro**

| Year | MDE[1] | No MDE[2] | AUD[3] | No AUD[4] | MDE and AUD[5] | No MDE or AUD[6] |
|---|---|---|---|---|---|---|
| 2005 | 18.05 [17.17-18.93] | 11.29 [10.77-11.8] | 55.13 [53.57-56.68] | 8.44 [8-8.88] | 58.13 [56.08-60.17] | 8.35 [7.94-8.77] |
| 2006 | 17.29 [16.43-18.15] | 10.77 [10.31-11.23] | 53.81 [52.23-55.38] | 8.04 [7.63-8.45] | 56.79 [54.55-59.03] | 7.94 [7.57-8.32] |
| 2007 | 17.45 [16.55-18.35] | 10.88 [10.39-11.37] | 54.2 [52.5-55.9] | 8.16 [7.7-8.61] | 56.92 [54.6-59.25] | 7.98 [7.57-8.4] |
| 2008 | 17.22 [16.27-18.17] | 10.73 [19.16-11.29] | 53.69 [51.87-55.51] | 8 [7.54-8.47] | 56.51 [54.11-58.9] | 7.86 [7.42-8.3] |
| 2009 | 16.48 [15.67-17.29] | 10.23 [9.76-10.7] | 52.28 [50.64-53.92] | 7.6 [7.21-7.98] | 55.12 [52.99-57.25] | 7.46 [7.09-7.84] |
| 2010 | 15.67 [14.83-16.51] | 9.69 [9.26-10.13] | 51.16 [49.54-52.77] | 7.29 [6.86-7.71] | 53.78 [51.68-55.88] | 7.1 [6.73-7.47] |
| 2011 | 15.74 [14.88-16.61] | 9.74 [9.25-10.23] | 51.92 [50.09-53.75] | 7.5 [7.05-7.94] | 54.3 [51.68-56.93] | 7.24 [6.82-7.66] |
| 2012 | 15.37 [14.6-16.14] | 9.5 [9.05-9.94] | 50.8 [49.23-52.36] | 7.19 [6.81-7.57] | 53.35 [51.21-55.48] | 6.98 [6.62-7.35] |
| 2013 | 15.34 [14.44-16.24] | 9.47 [8.91-10.04] | 50.75 [48.71-52.78] | 7.18 [6.72-7.63] | 53.21 [50.56-55.87] | 6.95 [6.49-7.4] |
| 2014 | 14.29 [13.48-15.11] | 8.78 [8.35-9.22] | 48.87 [47.28-50.46] | 6.69 [6.32-7.07] | 51.37 [49.02-53.71] | 6.49 [6.14-6.83] |
| 2016 | 10.26 [9.74-10.78] | 6.2 [5.91-6.48] | 39.28 [37.88-40.67] | 4.63 [4.39-4.87] | 41.64 [39.59-43.68] | 4.47 [4.26-4.69] |
| 2017 | 10.5 [9.95-11.04] | 6.34 [6.04-6.65] | 40.47 [38.02-41.92] | 4.85 [4.61-5.09] | 42.09 [39.97-44.2] | 4.55 [4.32-4.78] |
| 2018 | 9.78 [9.15-10.41] | 5.89 [5.56-6.23] | 38.35 [36.62-49.97] | 4.46 [4.19-4.73] | 40.05 [37.6-42.49] | 4.2 [3.95-4.45] |
| 2019 | 9.41 [8.75-10.07] | 5.66 [5.29-6.03] | 37.54 [35.74-39.35] | 4.32 [4.01-4.62] | 38.55 [36.12-40.99] | 3.96 [3.67-4.24] |

[1] MDE: AAPC = -5.0 (95% CI, -6.3 to -3.7), $p < 0.001$

[2] No MDE: AAPC = -5.3 (95% CI, -6.7 to -3.9), $p < 0.001$

[3] AUD: AAPC = -3.1 (95% CI, -3.7 to -2.5), $p < 0.001$

[4] No AUD: AAPC = -5.2 (95% CI, -6.7 to -3.6), $p < 0.001$

[5] MDE and AUD: AAPC = -3.1 (95% CI, -3.8 to -2.4), $p < 0.001$

[6] No MDE or AUD: AAPC = -5.6 (95% CI, -7.0 to -4.2), $p < 0.001$

**B. Small metro**

| Year | MDE[1] | No MDE[2] | AUD[3] | No AUD[4] | MDE and AUD[5] | No MDE or AUD[6] |
|---|---|---|---|---|---|---|
| 2005 | 18.7 [17.77-19.64] | 11.73 [11.19-12.26] | 56.49 [54.98-58] | 8.88 [8.43-9.32] | 59.36 [57.39-61.33] | 8.75 [8.33-9.17] |
| 2006 | 17.92 [16.94-18.9] | 11.2 [10.66-11.73] | 55.18 [53.6-56.76] | 8.46 [8.03-8.89] | 58.04 [55.8-60.27] | 8.33 [7.91-8.74] |
| 2007 | 18.09 [17.09-19.08] | 11.31 [10.77-11.86] | 55.57 [53.79-57.35] | 8.58 [8.07-9.09] | 58.17 [55.85-60.48] | 8.37 [7.91-8.82] |
| 2008 | 17.85 [16.87-18.83] | 11.15 [10.59-11.71] | 55.06 [53.35-56.78] | 8.42 [7.97-8.87] | 57.75 [55.45-60.06] | 8.24 [7.8-8.69] |
| 2009 | 17.08 [16.17-18] | 10.63 [10.11-11.16] | 53.66 [51.85-55.46] | 7.99 [7.53-8.46] | 56.37 [54.18-58.57] | 7.82 [7.38-8.26] |
| 2010 | 16.25 [15.28-17.23] | 10.08 [9.55-10.6] | 52.54 [50.77-54.31] | 7.67 [7.18-8.16] | 55.04 [52.87-57.22] | 7.44 [7.01-7.87] |
| 2011 | 16.33 [15.43-17.23] | 10.13 [9.63-10.63] | 53.3 [51.5-55.1] | 7.89 [7.43-8.35] | 55.57 [53.01-58.12] | 7.59 [7.16-8.01] |
| 2012 | 15.95 [15.15-16.74] | 0.99 [9.43-10.32] | 52.18 [50.73-53.63] | 7.57 [7.2-7.93] | 54.61 [52.58-56.65] | 7.32 [6.97-7.68] |
| 2013 | 15.91 [14.95-16.87] | 9.85 [9.26-10.45] | 52.13 [50.07-54.18] | 7.55 [7.07-8.04] | 54.48 [51.86-57.11] | 7.29 [6.81-7.77] |
| 2014 | 14.83 [13.96-15.7] | 9.14 [8.69-9.59] | 50.25 [48.66-51.85] | 7.05 [6.65-7.44] | 52.64 [50.32-54.96] | 6.8 [6.44-7.17] |
| 2016 | 10.67 [10.1-11.23] | 6.45 [6.15-6.75] | 40.6 [39.19-42.02] | 4.88 [4.62-5.13] | 42.88 [40.83-44.93] | 4.7 [4.46-4.93] |
| 2017 | 10.91 [10.31-11.51] | 6.61 [6.28-6.94] | 41.81 [40.29-43.33] | 5.12 [4.85-5.38] | 43.33 [41.19-45.48] | 4.78 [4.53-5.03] |
| 2018 | 10.17 [9.5-10.83] | 6.14 [5.79-6.49] | 39.66 [37.9-41.42] | 4.7 [4.41-4.99] | 41.28 [38.82-43.73] | 4.41 [4.15-4.68] |
| 2019 | 9.79 [9.09-10.48] | 5.9 [5.51-6.28] | 38.85 [37.02-40.67] | 4.55 [4.23-4.87] | 39.77 [37.33-42.21] | 4.16 [3.85-4.46] |

[1] MDE: AAPC = -5.0 (95% CI, -6.3 to -3.7), $p < 0.001$

[2] No MDE: AAPC = -5.0 (95% CI, -6.4 to -3.5), $p < 0.001$

[3] AUD: AAPC = -2.9 (95% CI, -3.7 to -2.1), $p < 0.001$

[4] No AUD: AAPC = -5.2 (95% CI, -6.6 to -3.6), $p < 0.001$

[5] MDE and AUD: AAPC = -3.0 (95% CI, -3.7 to -2.3), $p < 0.001$

[6] No MDE or AUD: AAPC = -5.6 (95% CI, -6.9 to -4.2), $p < 0.001$

**C. Non-metro**

| Year | MDE[1] | No MDE[2] | AUD[3] | No AUD[4] | MDE and AUD[5] | No MDE or AUD[6] |
|---|---|---|---|---|---|---|
| 2005 | 17.15 [16.2-18.09] | 10.68 [10.09-11.27] | 54.32 [52.51-56.12] | 8.19 [7.68-8.7] | 57.06 [54.89-59.24] | 8.03 [7.55-8.51] |
| 2006 | 16.42 [15.46-17.37] | 10.19 [9.63-10.75] | 52.99 [51.19-54.79] | 7.8 [7.33-8.27] | 55.72 [53.33-58.1] | 7.63 [7.18-8.09] |
| 2007 | 16.57 [15.63-17.52] | 10.29 [9.74-10.84] | 53.39 [51.45-55.32] | 7.91 [7.4-8.43] | 55.85 [53.41-58.29] | 7.67 [7.2-8.14] |
| 2008 | 16.35 [15.48-17.22] | 10.14 [9.62-10.67] | 52.88 [51.07-54.68] | 7.77 [7.32-8.21] | 55.43 [53.09-57.78] | 7.55 [7.13-7.97] |
| 2009 | 15.64 [14.84-16.43] | 9.67 [9.19-10.15] | 51.46 [49.74-53.18] | 7.37 [6.97-7.77] | 54.04 [51.91-56.16] | 7.17 [6.78-7.55] |
| 2010 | 14.86 [13.95-15.78] | 9.16 [8.64-9.69] | 50.34 [48.44-52.23] | 7.07 [6.58-7.55] | 52.7 [50.43-54.96] | 6.82 [6.38-7.26] |
| 2011 | 14.93 [14.08-15.79] | 9.21 [8.7-9.71] | 51.1 [49.27-52.93] | 7.27 [6.84-7.7] | 53.22 [50.6-55.84] | 6.95 [6.53-7.38] |
| 2012 | 14.58 [13.81-15.35] | 8.97 [8.51-9.44] | 49.98 [48.26-51.69] | 6.97 [6.56-7.39] | 52.26 [50.07-54.45] | 6.71 [6.31-7.1] |
| 2013 | 14.55 [13.75-15.35] | 8.95 [8.44-9.46] | 49.93 [47.93-51.92] | 6.96 [6.53-7.39] | 52.13 [49.59-54.66] | 6.67 [6.26-7.09] |
| 2014 | 13.55 [12.72-14.38] | 8.3 [7.84-8.76] | 48.05 [46.29-49.81] | 6.49 [6.08-6.9] | 50.28 [47.86-52.7] | 6.23 [5.84-6.61] |
| 2016 | 9.7 [9.15-10.26] | 5.84 [5.52-6.17] | 38.5 [36.89-40.11] | 4.49 [4.21-4.76] | 40.58 [38.45-42.72] | 4.29 [4.04-4.55] |
| 2017 | 9.93 [9.36-10.49] | 5.98 [5.65-6.32] | 39.68 [38.07-41.3] | 4.7 [4.44-4.97] | 41.03 [38.86-43.2] | 4.37 [4.11-4.63] |
| 2018 | 9.24 [8.61-9.87] | 5.56 [5.2-5.91] | 37.57 [35.66-39.49] | 4.32 [4.02-4.62] | 39 [36.5-41.51] | 4.03 [3.75-4.31] |
| 2019 | 8.89 [8.27-9.51] | 5.34 [4.98-5.69] | 36.78 [34.95-38.6] | 4.18 [3.88-4.49] | 37.53 [35.14-39.91] | 3.79 [3.51-4.08] |

[1] MDE: AAPC = -5.0 (95% CI, -6.3 to -3.7), $p < 0.001$

[2] No MDE: AAPC = -5.3 (95% CI, -6.6 to -3.9), $p < 0.001$

[3] AUD: AAPC = -3.0 (95% CI, -2.2 to -6.9), $p < 0.001$

[4] No AUD: AAPC = -5.1 (95% CI, -6.6 to -3.6), $p < 0.001$

[5] MDE and AUD: AAPC = -3.1 (95% CI, -3.8 to -2.4), $p < 0.001$

[6] No MDE or AUD: AAPC = -5.6 (95% CI, -6.9 to -4.2), $p < 0.001$
